# Intravenous corticosteroid treatment in adult patients with sepsis defined by the Sepsis-3 criteria: a systematic review and meta-analysis

**DOI:** 10.1101/2021.10.17.21265100

**Authors:** Yu-Pu Wu, Cheng-Kuan Lin, Rikuta Hamaya, Fei-Yang Huang, Yung-Shin Chien, Yu-Tien Hsu, Szu-Ta Chen, Stefania Papatheodorou

**Affiliations:** Department of Epidemiology, Harvard T.H. Chan School of Public Health, Boston, MA, U.S.A.; Department of Environmental Health, Harvard T.H. Chan School of Public Health, Boston, MA, U.S.A.; School of Arts and Sciences, Massachusetts College of Pharmacy and Health Sciences, MA, U.S.A.; Department of Physiology, Development and Neuroscience, University of Cambridge, United Kingdom; Department of Social and Behavioral Sciences, Harvard T.H. Chan School of Public Health, Boston, MA, U.S.A.; Population Health Sciences, Graduate School of Arts and Sciences, Harvard University, U.S.A.

**Keywords:** Sepsis, Shock, Septic, Survival, Glucocorticoids, Mortality

## Abstract

**Objectives:** To summarize the effects of intravenous corticosteroid treatment for sepsis defined by the Sepsis-3 criteria in adult patients.

**Design:** Systematic review and meta-analysis.

**Methods:** We searched RCTs from PubMed, Embase, ClinicalTrials.gov, Cochrane Central Register of Controlled Trials, Web of Science, and International Clinical Trials Registry Platform from inception to July 12th, 2019 and updated on June 28th, 2020. Conference proceedings from relevant societies and the reference lists of previous reviews were manually screened. Abstract or full-text articles were screened by two independent investigators. We included RCTs where (1) the participants had infections and the baseline Sequential Organ Failure Assessment (SOFA) score ≥ 2 (the Sepsis-3 definitions) (2) the intervention involved any intravenous corticosteroids; (3) the control group received placebo or standard of care (4) the outcomes of interest included mortality or clinical recovery. We chose the 28-day mortality as the pre-specified primary outcome and risk ratio (RR) as the effect measure. We followed PRISMA guidelines and chose random-effects models for the pooled analyses.

**Results:** This study included 24 RCTs and 19 of them (7,115 participants) reported the 28-day mortality. Pooled analyses showed that intravenous corticosteroid treatment compared to placebo or standard of care was not associated with a lower risk of 28-day mortality (RR, 0.88; 95%CI, 0.73 to 1.05), but with a higher risk of hyperglycemia (RR, 1.16; 95%CI, 1.06 to 1.27). Sensitivity analysis of high-quality studies revealed a similar result for the 28-day mortality (RR, 0.95; 95%CI, 0.86 to 1.05).

**Conclusions:** Our findings suggested that intravenous corticosteroids compared to placebo or standard of care may not reduce the 28-day mortality in adult patients with sepsis defined by the Sepsis-3 criteria. Further studies are warranted to clarify the roles of disease severity and treatment timing in the effects of corticosteroid treatment in this population.

**PROSPERO registration number:** CRD42019143083

**Strengths and limitations of this study:**

- This is the first systematic review and meta-analysis that summarized the effects of intravenous corticosteroid treatments in patients with sepsis defined by the Sepsis-3 criteria.
- We provide the quality of evidence to support the development of treatment guidelines specific to the Sepsis-3 cohort.
- We only include randomized controlled trials in this systematic review and meta-analysis, which exclude less controlled evidences from clinical settings closer to our daily practice.
- The Sepsis-3 definitions will be retrospectively applied to the included studies, so clinical trials without enough reported baseline data available may be excluded.

## Introduction

Controversy over the use of intravenous corticosteroid in patients with sepsis or septic shock remained unsettled for decades.^1^ Published randomized controlled trials (RCTs) reported inconsistent findings.^1^ In particular, the two largest RCTs, the APROCCHSS trial^2^ and the ADRENAL trial,^3^ published in 2018 did not agree with each other. The APROCCHSS trial suggested that corticosteroid use was associated with reduced 90-day mortality,^2^ whereas the ADRENAL trial did not.^3^ An explanation for this inconsistency could be different inclusion criteria for patients with septic shock between these two studies. The ADRENAL trial^3^ defined the study cohort using an earlier definition for sepsis,^4^ the systematic inflammatory response syndrome (SIRS) following infection. Instead, the APROCCHSS trial enrolled participants based on the Sequential Organ Failure Assessment (SOFA) score.^2^ As a result, the APROCCHSS trial had a higher mortality rate in the control group compared to that in the ADRENAL trial (49.1% vs 28.8%).^2 3^ Additional inconsistencies of the inclusion criteria also existed among other RCTs. For example, some trials enrolled patients with concomitant use of vasopressor agents,^5-7^ whereas others did not.^8-10^ Due to inconsistent definitions of patients with sepsis, the results are difficult to summarize to inform clinical practice.

In 2016, a new consensus definition for the diagnosis of sepsis, The Third International Consensus Definitions for Sepsis and Septic Shock (Sepsis-3), was proposed based on the SOFA score to standardize and operationalize the diagnostic criteria for sepsis and septic shock.^11^ The use of the SOFA score not only remedied the high false positive rate of the SIRS definition in identifying patients with sepsis,^12^ but was validated as a better predictor for mortality.^11^ Thus, the Sepsis-3 definitions were increasingly adopted by researchers and clinicians to ensure consistency in clinical studies and daily practice.^11 13-15^

After the Sepsis-3 definitions were proposed, to the best of our knowledge, only one post-hoc analysis of the ADRENAL trial has applied the Sepsis-3 criteria for septic shock,^16^ but the association between intravenous corticosteroids and mortality in patients with sepsis defined by the Sepsis-3 criteria has not yet been summarized. Thus, we aimed to conduct this systematic review and meta-analysis to summarize the efficacy of corticosteroids in patients with sepsis defined by the Sepsis-3 criteria.

## Methods

This systematic review and meta-analysis was conducted based on a pre-specified protocol registered in the International prospective register of systematic reviews (PROSPERO) on October 24^th^, 2019 (registered number: CRD42019143083). We followed the PRISMA reporting guideline^17^ to report our study design and findings.

### Search strategy

We searched published or completed RCTs on the efficacy of corticosteroids in adult patients with sepsis by using the searching terms: (1) corticosteroids; (2) infectious diseases or syndromes associated with sepsis; and (3) randomized controlled trials and their synonyms. The searching included multiple databases (i.e. PubMed, Embase, Cochrane Central Register of Controlled Trials, Web of Science, ClinicalTrials.gov, and International Clinical Trials Registry Platform) as well as online resources of leading societies for intensive care, such as Society of Critical Care Medicine, International Sepsis Forum, European Society of Intensive Care Medicine, American Thoracic Society, and World Federation of Societies of Intensive and Critical Care. All the above-mentioned sources were searched from the inception till July 12, 2019 and updated on June 28th, 2020. Reference lists of the previous relevant review articles^18-20^ were further screened manually. The detailed search strategy was reported in eTable 1 (see online supplementary materials).

### Eligibility criteria

PICOS statement (Patients/ Intervention/ Comparison/ Outcome/ Study design) were set up *a priori* as below: (1) Adult patients (age ≥18 years) that had infections and baseline SOFA score ≥2, as defined by the Sepsis-3 criteria for sepsis. The baseline SOFA score was either directly reported in the trials, or indirectly derived from the baseline biochemical laboratory data and the inclusion criteria of the trials. If the baseline SOFA score was reported, we included the trials when approximately 97.5% of subjects fulfilled the Sepsis-3 criteria for sepsis, indicated by the approximation that the mean of baseline SOFA score minus twice of the standard deviation (SD) was equal or larger than 2. If the median with interquartile range (IQR) was reported, we approximated the mean with the median, and assumed the IQR to be 1.35 SD. The SOFA scores were calculated based on the Sepsis-3 criteria^11^; (2) Interventions included any intravenous corticosteroids. (3) Comparison group received either placebo or standard of care, such as antibiotics, intravenous fluid support, mechanical ventilation, and vasopressors. (4) Studies reported at least one of the following pre-specified outcomes were eligible: 28-day mortality (our primary outcome of interest), 90-day mortality, length of intensive care unit (ICU) stay, length of hospital stay, SOFA score on day 7, in-hospital mortality, ICU mortality, and adverse events including hyperglycemia, gastrointestinal bleeding, superinfection, and the total numbers of any adverse events; (5) We limited our search in randomized controlled trials. Therefore, studies in which the treatments were not randomly assigned were excluded.

To focus on the efficacy of corticosteroids, we excluded trials in which both the experimental group and the control group received corticosteroid treatment. Notably, abstract-only studies that fulfilled our PICOS statement were still eligible for our study. No language restriction was applied.

### Study selection

Each article was independently screened, assessed and selected by at least two authors (Y.W., C.L., F.H., Y.C., Y.H., S.C.). In case of disagreements, consensus was reached through discussion among the investigators or after consultation with a third reviewer (S.P.).

### Data extraction

Four independent reviewers (Y.W., R.H., F.H., Y.C.) extracted relevant information from the included trials. Specifically, we extracted data including general characteristics of the articles (e.g. authors, years of publication, sample sizes), information related to the baseline SOFA scores (e.g. the inclusion criteria or the biochemical laboratory data), and information of the treatment (types of medications, dosages, and treatment durations), and all outcomes of interest. All disagreements were resolved through the discussion among reviewers. Missing or unpublished data were obtained by consulting previous related review articles.^18 19^

### Risk of bias and quality of evidence

Two reviewers (Y.W. and R.H.) independently assessed the risk of bias for each included trial using the revised Cochrane risk-of-bias tool, RoB2.^21^ The same reviewers also independently adopted the Grading of Recommendations Assessment, Development and Evaluation (GRADE) approach to evaluate the quality of evidence for each outcome of interest.^22^ The third reviewer (S.P.) was consulted in case of disagreements.

Specifically, there were five domains in the RoB2, including the randomization process, deviations from the intended interventions, the missing outcome data, the measurement of the outcome, and the selection of the reported results. Each domain was judged as ‘low’ or ‘high’ risk of bias, or with ‘some concerns’. An overall risk of bias was reported for each trial, and a trial was considered having some concerns or high-risk for overall bias if judged as ‘some concerns’ or ‘high’ risk of bias in any of the five domains, respectively. As recommended by the GRADE system, the quality of evidence for each outcome of interest was evaluated based on the risk of bias,^23^ imprecision,^24^ inconsistency,^25^ indirectness,^26^ and the reporting bias.^27^ An overall quality of evidence for each outcome was rated as high, moderate, low, or very low.

### Data synthesis

We conducted pooled analyses for all the outcomes of interest using random-effects models with Hartung-Knapp-Sidik-Jonkman (HKSJ) method which outperformed the DerSimonian-Laird method particularly in having lower type-I error rates when sample sizes varied across trials.^28^ Risk ratio (RR) of dichotomous outcomes and risk difference (RD) of continuous outcomes were reported with 95% confidence intervals (95% CI) using Mantel-Haenszel method and inverse variance method, respectively. Statistical significance level was defined as a two-tailed P-value <0.05. We used Chi-square test and I^2^ statistics to evaluate the magnitude of the heterogeneity for each outcome. Significance level of the Chi-square test was defined as a two-tailed P-value <0.1. As defined by the Cochrane Handbook,^29^ moderate or substantial heterogeneity was considered if I^2^ statistics ≥30% or ≥50%, respectively. The reporting bias for each outcome was visually evaluated based on the funnel plot. If at least 10 trials were pooled, the Egger test for the continuous outcomes or the Harbord test for the dichotomized outcomes were used to test the asymmetry of the funnel plot with the significance level of 0.1.^29 30^

### Subgroup analysis

We performed pre-specified subgroup analyses based on the duration of corticosteroid treatments (≤7 days vs >7 days), average daily hydrocortisone equivalent dose (≤200mg vs >200mg), severity of sepsis (sepsis vs septic shock based on the Sepsis-3 definitions), and types of the corticosteroids (hydrocortisone, methylprednisolone, hydrocortisone plus fludrocortisone, betamethasone, or dexamethasone). A mixed-effects model^31^ was used for the subgroup comparisons with a significance level of 0.05.

### Sensitivity analysis

We performed the sensitivity analyses by (1) excluding trials with < 100 total patients; (2) excluding studies with <10 events in either the experimental group or the control group; (3) excluding studies with some concerns or high risk of overall bias; (4) excluding studies without a double-blind design; (5) excluding studies published before 2012 because an updated international guideline for the treatment of sepsis and septic shock was published in 2012^32^; (6) using a random-effects model with DerSimonian-Laird method; and (7) using a fix-effects model.

R (version 3.6.1) and packages meta, robvis, and dmetar^33^ were used for all the analyses.

### Patient and public involvement

This systematic review and meta-analysis did not have direct patients or public involvement.

## Results

Of the total 21,143 screened articles, 24 RCTs^2 3 5-10 34-49^ fulfilled our inclusion criteria and were included for meta-analysis (Figure 1). Evidence that the included subjects fulfilled the Sepsis-3 criteria (baseline SOFA score≥2) were reported in Table 1. Of the 24 included trials, 1 trial enrolled participants based on the Sepsis-3 criteria^49^; and 9 trials^2 5 8-10 34 37 38 40^ directly reported the patients’ baseline SOFA scores. We indirectly estimated the baseline SOFA scores for the other 14 studies^3 6 7 35 36 39 41-48^ based on the reported biochemical laboratory data or the inclusion criteria of these trials. Notably, 2 RCTs^45 48^ were evaluated only by the abstracts due to the inaccessibility of the main texts. For the treatment group, hydrocortisone,^3 6 8-10 34 36-41 43 45-47^ methylprednisolone,^35 44^ hydrocortisone plus fludrocortisone,^2 7^ and dexamethasone^5^ were used in 16, 2, 2, and 1 trials, respectively. Two other studies used the combination of hydrocortisone, Vitamin C, and thiamine,^48 49^ and the other trial administered either dexamethasone or methylprednisolone to the participants in the treatment group.^42^ For the control group, 20 RCTs^2 3 5-10 34-40 42 43 45-47^ compared corticosteroid use with placebo, whereas 4 studies^41 44 48 49^ provided standard of care for the patients in the control group.

**Table 1.** Characteristics of the 24 Included Trials

| Trial | Patients, No. (Sites, No.) | Justification of Sepsis (SOFA score $\geq 2$) | Intervention | Comparator | Primary Outcome |
| --- | --- | --- | --- | --- | --- |
| CSG 1963<sup>46</sup> | 194 (5) | All patients required vasopressors | HC 300mg for 1 day then tapered gradually for 6 days | Placebo | In-hospital mortality |
| Sprung 1984<sup>42</sup> | 59 (2) | PaO<sub>2</sub>/FiO<sub>2</sub> <300 and 93% of patients required vasopressors | One dose of MPDN 30mg/kg, or DM 6mg/kg; if shock persisted, the same dose was given again | Placebo | Hospital mortality |
| Bollaert 1998<sup>39</sup> | 41 (2) | All patients required vasopressors | HC 100mg/8h with taper dose until shock reversal with additional days for taper dose | Placebo | Shock reversal at day 7 |
| Chawla 1999<sup>45,a</sup> | 44 (1) | All patients required vasopressors | HC 100mg/8h for 3 days, then tapered for four days | Placebo | Cessation of shock |
| Briegel 1999<sup>40</sup> | 40 (1) | Baseline SOFA score of both groups:<sup>b</sup> 12.8 (0.6); 11.8 (0.6) | HC 100mg loading dose followed by 0.18mg/kg/h with taper dose until the cessation of vasopressor | Placebo | Time to cessation of vasopressor support |
| Annane 2002<sup>7</sup> | 300 (19) | All patients required vasopressors | HC (50mg/6h) and FC (oral 50mcg/d) for 7 days | Placebo | 28-day mortality |
| Oppert 2005<sup>10</sup> | 41 (1) | Baseline SOFA score of both groups:<sup>c</sup> 11 (10 to 13); 10 (8.5 to 13) | HC 50mg loading dose followed by 0.18mg/kg/h with taper dose until shock reversal | Placebo | Time to cessation of vasopressor support |
| Confalonieri 2005<sup>36</sup> | 46 (6) | PaO<sub>2</sub>/FiO<sub>2</sub> <300 | HC 200mg loading dose followed by 10mg/h for 7 days | Placebo | Improvement in PaO<sub>2</sub>:FiO<sub>2</sub> by day 8 |
| Cicarelli 2007<sup>5</sup> | 29 (1) | Baseline SOFA score of both groups:<sup>b</sup> 10 (2); 9 (3) | 3 doses of DM 0.2mg/kg at intervals of 36 h | Placebo | 7-day mortality |
| Sprung 2008<sup>9</sup> | 499 (52) | Baseline SOFA score of both groups:<sup>b</sup> 10.6 (3.4); 10.6 (3.2) | HC 50mg/6h for 5 days + 50mg/12h for 3 days + 50mg/d for 3 days | Placebo | 28-day mortality |
| Hu 2009<sup>41</sup> | 77 (1) | All patients required vasopressors | HC 50mg/6h for 7 days + 50mg/8h for 3 days + 50mg/12h for 2 days + 50mg/d for 2 days | Standard of care | Lactate clearance and the usage of norepinephrine |
| Arabi 2010<sup>38</sup> | 75 (1) | Baseline SOFA score of both groups:<sup>b</sup> 14.6 (3.7); 14.3 (3.7) | HC 50mg/6h with taper dose until shock reversal | Placebo | 28-day mortality |
| Sabry 2011<sup>34</sup> | 80 (3) | Baseline SOFA score of both groups:<sup>b</sup> 8.2 (1.46); 8.5 (1.52) | HC 200 mg loading dose followed by 12.5mg/h for 7 days | Placebo | Mean SOFA score by day 8 |
| Fernández-Serrano 2011<sup>35</sup> | 56 (1) | PaO<sub>2</sub>/FiO<sub>2</sub> <300 | MPDN 200mg bolus dose followed by 20mg/6h 3 days, 20mg/12h 3 days, and 20mg/d 3 days | Placebo | Respiratory failure requiring ventilation |

Table 1. Characteristics of the 24 Included Trials (continued)
| Trial | Patients, No.<br>(Sites, No.) | Justification of Sepsis<br>(SOFA score $\geq 2$) | Intervention | Comparator | Primary Outcome |
| --- | --- | --- | --- | --- | --- |
| Gordon<br>2014<sup>43</sup> | 61 (4) | All patients required<br>vasopressors | HC 50mg/6h for 5 days +<br>50mg/12h for 3 days +<br>50mg/d for 3 days | Placebo | difference in plasma<br>vasopressin<br>concentrations<br>between groups |
| Gordon<br>2016<sup>6</sup> | 409 (18) | All patients required<br>vasopressors | HC 50mg/6h for 5 days +<br>50mg/12h for 3 days +<br>50mg/d for 3 days | Placebo | Kidney failure free<br>days to day 28 |
| Tongyoo<br>2016<sup>37</sup> | 197 (1) | Baseline SOFA score<br>of both groups:<sup>b</sup><br>10.9 (3.5); 10.8 (3.6) | HC 50mg/6h for 7 days | Placebo | 28-day mortality |
| Li<br>2016<sup>44</sup> | 58 (1) | All patients required<br>vasopressors | MPDN 80mg/d for 7 days | Standard of<br>care | Changes in<br>PaO<sub>2</sub>:FiO<sub>2</sub> and<br>CRP by day 1, 4,<br>and 8 |
| Lv<br>2017<sup>8</sup> | 118 (1) | Baseline SOFA score<br>of both groups:<sup>b</sup><br>11.9 (3.3); 9.9 (3.0) | HC 200mg/d for 6 days<br>with taper dose | Placebo | 28-day mortality |
| Annane<br>2018<sup>2</sup> | 1241 (34) | Baseline SOFA<br>score:<sup>b</sup> 12 (3) | HC 50mg/6h + oral FC<br>50mcg/d for 7 days | Placebo | 90-day mortality |
| Doluee<br>2018<sup>47</sup> | 160 (1) | Not respond to<br>vasopressors | HC 50mg/6h for 7 days | Placebo | 28-day all-cause<br>mortality |
| Venkatesh<br>2018<sup>3</sup> | 3800 (69) | All patients received<br>vasopressors | HC 200mg/d, maximum<br>of 7 days | Placebo | 90-day mortality |
| Fan<br>2019<sup>48,a</sup> | 18 (1) | All patients received<br>vasopressors | HC 50mg/6h for 7 days or<br>until ICU discharge<br>combined with Vitamin C<br>and thiamine<sup>d</sup> | Standard of<br>care | 28-day mortality |
| Wani<br>2020<sup>49</sup> | 100 (1) | Based on the Sepsis-3<br>criteria for sepsis;<br>Baseline SOFA score<br>of both groups:<sup>b</sup><br>9.36 (3.66); 9.22<br>(3.54) | HC 50mg/6h for 7 days or<br>until ICU discharge with<br>taper dose for 3 days<br>combined with Vitamin C<br>and thiamine<sup>e</sup> | Standard of<br>care | In-hospital mortality |

Abbreviation: HC, hydrocortisone; MPDN, methylprednisolone; FC, fludrocortisone; DM, dexamethasone; MAP, Mean arterial pressure; CRP, C-reactive protein; ICU, intensive care unit;
<sup>a</sup>Abstract only;
<sup>b</sup>Mean (SD);
<sup>c</sup>Median (IQR);
<sup>d</sup>Intravenous Vitamin C 1.5g/6h for 4 days/until ICU discharge + intravenous thiamine 200mg/12h for 4 days/ until ICU discharge;
<sup>e</sup>Intravenous Vitamin C 1.5g/6h for 4 days/until hospital discharge + intravenous thiamine 200mg/12h for 4 days/ until hospital discharge.

**Figure 1.**
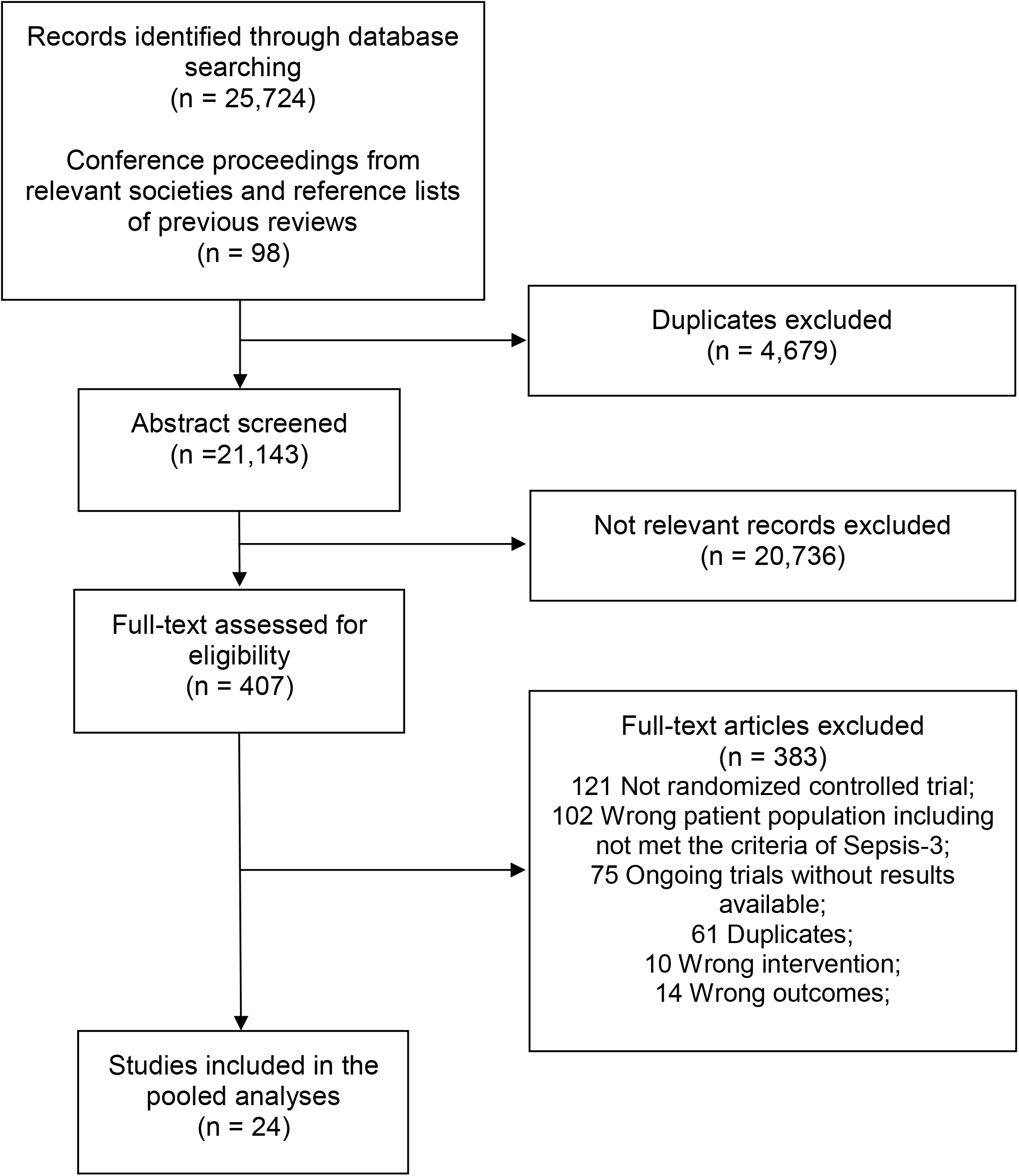
Study Selection Process with Reasons for Exclusion

### Risk of bias and quality of evidence

eFigure 1 and 2 (see online supplementary materials) displays the risk of bias for each domain. Of the 24 trials, 6 trials^3 6 7 9 37 47^ were considered having low risk of bias across all 5 domains, 8 studies^2 5 8 34 36 40 44 45^ had some concerns, and 10 studies^10 35 38 39 41-43 46 48 49^ had overall high risk of bias. Trials with overall high risk of bias were mostly due to non-double blinded designs^42-44 48 49^ or premature termination.^38 39^

eTable 2 (see online supplementary materials) details the judgement for the risk of bias. The GRADE assessment for each outcome is shown in Table 2. An overall quality of evidence for the 28-day mortality was high, and most secondary outcomes had medium-to-high quality.

**Table 2.** Summary of Findings with Quality of Evidence

| Outcomes | Patients, No.<br>(Studies, No.) | Point estimate,<br>RR, or MD<br>(95% CI) | Absolute effect estimates,<br>Control vs Treated with 95% CI,<br>per 1000 individuals | P-value | I<sup>2</sup><br>(%) | Quality<br>(GRADE) |
| --- | --- | --- | --- | --- | --- | --- |
| Primary outcome |  |  |  |  |  |  |
| 28-day mortality | 7115 (19) | 0.88<br>(0.73, 1.05) | 318 vs 280 (232 to 334) | 0.15 | 20 | High |
| Secondary outcomes |  |  |  |  |  |  |
| 90-day mortality | 5136 (4) | 0.92<br>(0.84, 1.01) | 342 vs 315 (287 to 345) | 0.07 | 0 | Medium<sup>c</sup> |
| In-hospital mortality | 7042 (17) | 0.94<br>(0.82, 1.08) | 343 vs 322 (281 to 370) | 0.37 | 0 | High |
| ICU mortality | 6622 (13) | 0.83<br>(0.67, 1.03) | 294 vs 244 (197 to 303) | 0.08 | 9 | Medium<sup>c</sup> |
| Length of hospital stay | 6751 (14) | -1.92<br>(-4.15, 0.30) | NA | 0.08 | 41 | Medium<sup>b</sup> |
| Length of ICU stay | 6721 (14) | -1.43<br>(-3.14, 0.28) | NA | 0.09 | 51 | Medium<sup>b</sup> |
| SOFA score at day 7 | 1938 (8) | -1.37<br>(-1.91, -0.82) | NA | <0.01<sup>*</sup> | 56 | Medium<sup>b</sup> |
| Adverse events |  |  |  |  |  |  |
| Hyperglycemia | 6144 (10) | 1.16<br>(1.06, 1.27) | 284 vs 329 (301 to 361) | <0.01<sup>*</sup> | 0 | Medium<sup>c</sup> |
| GI bleeding | 6462 (16) | 1.10<br>(0.73, 1.65) | 27 vs 30 (20 to 45) | 0.61 | 0 | Medium<sup>a</sup> |
| Superinfection | 6368 (14) | 1.00<br>(0.78, 1.27) | 192 vs 192 (150 to 244) | 0.97 | 0 | High |
| Any adverse events | 5881 (9) | 1.07<br>(0.65, 1.77) | 164 vs 176 (107 to 290) | 0.77 | 65 | Low<sup>ab</sup> |
| Abbreviation: RR, Risk Ratio; MD, Mean Difference; NA, Not Applicable; GI, gastrointestinal.<br><sup>a</sup>Downgraded for imprecision because 95% CI of RR covered both 0.75 and 1.25;<br><sup>b</sup>Downgraded for inconsistency because of I<sup>2</sup> ≥ 30;<br><sup>c</sup>Downgraded for publication bias judged by visually inspecting the funnel plot or based on Egger/Harbord test;<br><sup>*</sup>Statistically significant |  |  |  |  |  |  |

### Primary outcome

Nineteen trials^2 3 5-10 36-40 42-45 47 48^ reported the risks of 28-day mortality in which 3,564 out of 7,115 participants (50.1%) received corticosteroid treatments. The summarized risk of 28-day mortality in patients with sepsis was not lower after corticosteroid treatment compared to the control group with placebo or standard of care (Figure 2: RR, 0.88; 95% CI, 0.73 to 1.05; P=0.15; I^2^ = 20%). No obvious asymmetry was found by visual inspections of the funnel plot (eFigure 3 in the online supplementary materials). The Harbord test also supported the absence of reporting bias (P=0.11). All the sensitivity analyses except the results from the fix-effects model were consistent with the primary analysis (Table 3 and eFigure 4-10 in the online supplementary materials). The sensitivity analysis based on 6 trials with low risk of overall bias agreed with the primary analysis (eFigure 6 in the online supplementary materials: RR, 0.95; 95% CI, 0.86 to 1.05).

**Figure 2.**
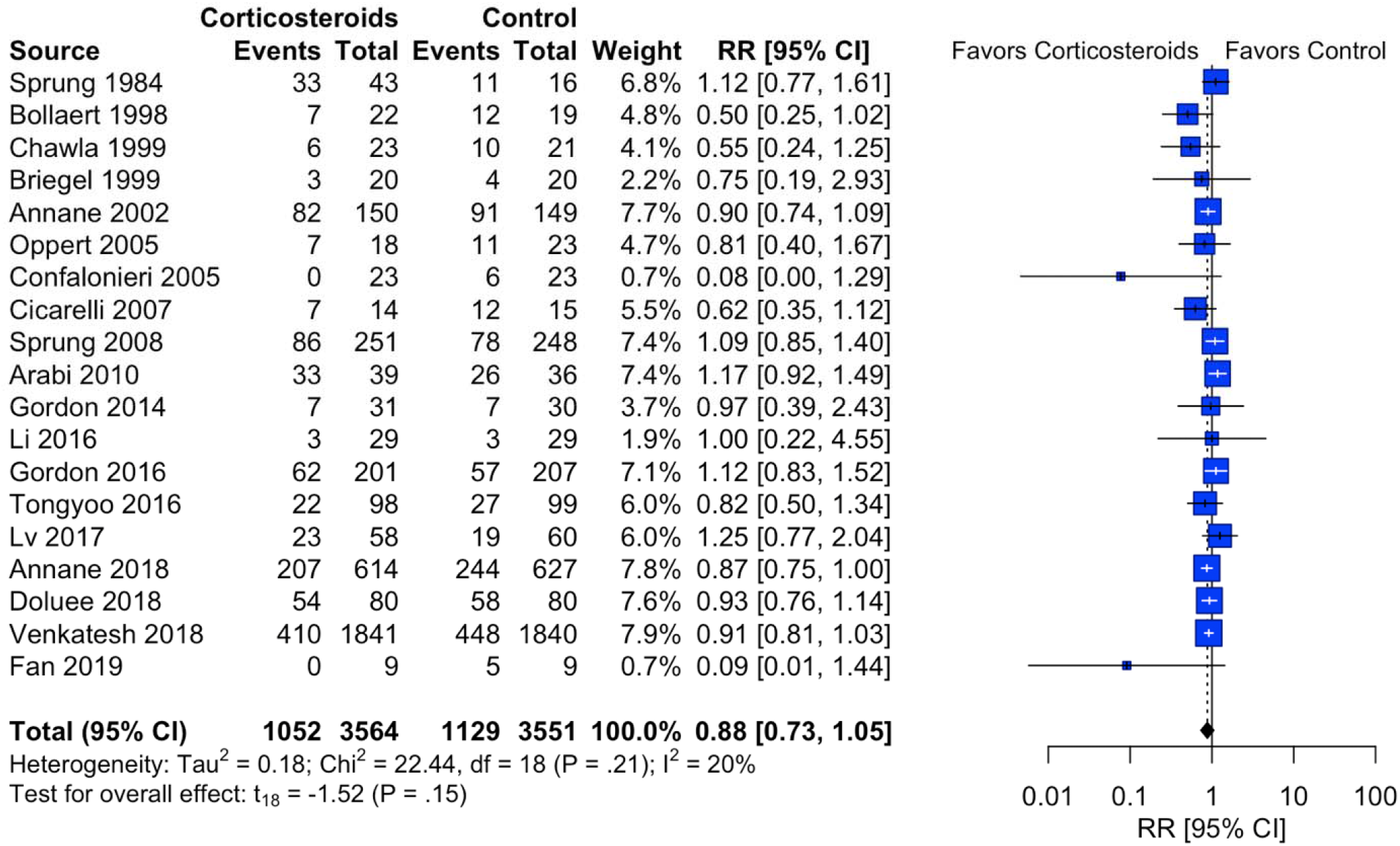
Forest Plot of the Pooled Effect of Corticosteroids Compared to Placebo or Standard of Care on 28-day Mortality in Patients with Sepsis Defined by the Sepsis-3 Criteria Abbreviation: RR, Risk Ratio; We used the random-effects model with Mantel-Haenszel method to estimate the risk ratios; Size of blue squares, weight of each trial; horizontal lines, 95% CIs; diamond, the overall risk ratio with 95% CI.

**Table 3.**
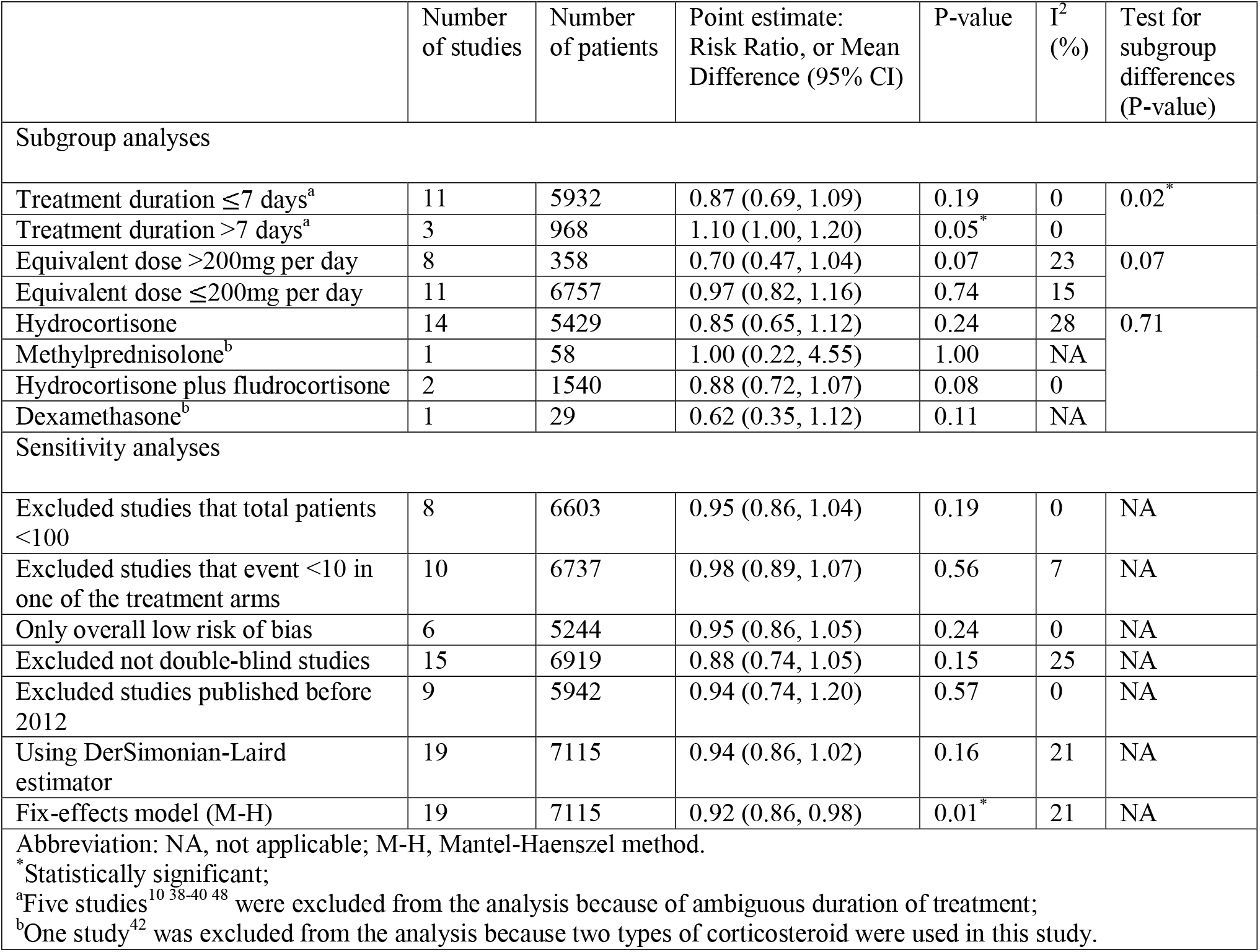
Subgroup and Sensitivity Analyses

### Secondary outcomes and adverse events

Table 2 and eFigure 11-20 in the online supplementary materials summarizes the pooled estimates and forest plots for all the secondary outcomes and adverse events, respectively. Intravenous corticosteroids were associated with lower SOFA scores on day 7 [mean differences (MD), −1.37; 95% CI = −1.91 to −0.81] but showed no benefits in mortality related outcomes and the length of hospital or ICU stay in patients with sepsis defined by the Sepsis-3 criteria. However, corticosteroid use was associated with higher risks of hyperglycemia (RR, 1.16; 95% CI, 1.06 to 1.27), summarized from 10 RCTs including more than 6,000 patients. The funnel plots (eFigure 21-30 in the online supplementary materials) suggest that the 90-day mortality, ICU mortality, and hyperglycemia suffered from potential reporting bias.

### Subgroup analysis

Subgroup analyses suggested that risks of mortality may vary with the durations of corticosteroid treatments: although patients with sepsis receiving corticosteroid treatments for more than 7 days had a borderline increased risk of mortality (RR, 1.10; 95% CI, 1.00 to 1.20), the risk remained the same for those with treatment duration less than 7 days (RR, 0.87; 95% CI, 0.69 to 1.09) (Table 3). Overall, there was no association between corticosteroid treatments and 28-day mortality, regardless of the types and dosages of corticosteroids. eFigure 31-41 (see online supplementary materials) displays the forest plots for the subgroup analyses. Importantly, we did not perform the subgroup analysis stratified based on disease severity (septic shock or not) because all included RCTs that reported the 28-day mortality had a mixed study cohort comprising patients with only sepsis and those who had progressed to septic shock (eTable 3 in the online supplementary materials).

## Discussion

In this systematic review and meta-analysis of 24 RCTs, we found that intravenous corticosteroids were not associated with lower 28-day mortality in adult patients with sepsis defined by the Sepsis-3 definitions, but were significantly associated with lower SOFA scores on day 7 and increased risks of hyperglycemia.

Based on extensive search in PubMed and Embase, we found 8 systematic reviews and meta-analyses^18-20 50-54^ on the use of corticosteroids in patients with sepsis had been published since 2018. Although all of these reviews included the two largest RCTs, the ADRENAL trial^3^ and the APROCCHSS trial,^2^ and included a substantial number of other RCTs, they reported contradictory results. In particular, 4 reviews^18 19 50 51^ reported that corticosteroids significantly reduced the risk of 28-day mortality, but other four studies^20 52-54^ found that intravenous corticosteroids did not lower the risk of either short-term or 28-day mortality. In addition to possible factors such as various outcome definitions or different modeling approaches, we argued that the mixed findings can only be clarified if we first properly standardized the inconsistent definitions for sepsis across previous trials. Our study overcame this limitation by following the Sepsis-3 criteria to define our study cohorts. This precise cohort definition allowed for accurate identification of patients with potential higher mortality rates and reduced the heterogeneity across the included trials, indicated by the I^2^ statistics <30 in most of our outcomes (Table 2).

However, our findings should be taken with caution because the disease severity (sepsis vs septic shock or high vs low SOFA score) seemed to modify the association between corticosteroid treatments and the 28-day mortality in the Sepsis-3 cohort. Although our analysis showed that corticosteroids were not associated with reduced 28-day mortality, the post hoc analysis of the ADRENAL trial^16^ showed that intravenous corticosteroid significantly reduced the 28-day mortality in patients who fulfilled the Sepsis-3 criteria for septic shock (odds ratio, 0.80; 95% CI, 0.64 to 0.99, p=0.04). In addition, the APROCCHSS trial,^2^ which included patients with mean baseline SOFA score of 12—much higher than the SOFA score≥2 required by the Sepsis-3 criteria for sepsis—showed that corticosteroids were associated with reduced 90-day mortality (RR, 0.88; 95% CI, 0.78 to 0.99, p=0.03). In our study, however, because many of the cohorts included patients from sepsis to septic shock and because not all RCTs reported baseline SOFA scores (Table 1 and eTable 3 in the online supplementary materials), we cannot accurately stratify the study cohorts according to the disease severity. Thus, more research based on well-defined patient inclusion criteria is needed to further clarify whether disease severity may modify the effects of corticosteroids in patients with sepsis.

In addition, our study showed that intravenous corticosteroids were associated with lower SOFA scores on day 7, but the effect did not sustain to improve 28-day survival. This paradoxical phenomenon could be explained by glucocorticoid resistance induced by exogenous corticosteroid, the altered corticosteroid metabolism, and biphasic immunological responses in critical illness. First, the administration of corticosteroid may induce glucocorticoid resistance,^55 56^ which compromised the efficacy of corticosteroids. In addition, reduced cortisol metabolism increases cortisol levels in patients with critical illness,^57^ which was associated with higher mortality in patients with severe sepsis.^58^ Furthermore, biphasic immunological states throughout the clinical course of sepsis,^59 60^ including an early inflammatory response followed by a chronic immunosuppressed status. This implied that treatment timing may influence the association between corticosteroids and the 28-day mortality because corticosteroid treatments may provide short-term benefits in the early inflammatory phase but have long-term negative impacts in the immunosuppressed state.^59^ Our subgroup analysis showed that corticosteroid treatment > 7 days was associated with higher 28-day mortality also supported this argument based on the biphasic immunological responses. This subgroup analysis, however, included only 3 studies^6 9 43^ and therefore required further research to confirm the influences of treatment timing on the efficacy of corticosteroids.

### Clinical implications

The recently updated Surviving Sepsis Campaign guidelines^61^ only recommend the use of intravenous corticosteroid for patients with septic shock who remain hemodynamically unstable after fluid resuscitation and vasopressor support. However, this recommendation was based on the evidence established from studies where participants were defined by the previous definitions.^61^ The findings of this meta-analysis can, therefore, bridge the clinical gap between guideline recommendations and current evidence to refine clinical use of corticosteroids in patients with sepsis defined by the Sepsis-3 criteria.

### Limitations

This study has several limitations. First, although small study effect, reporting bias, and chance may bias our findings, we confirmed that the results in the sensitivity analysis after excluding small studies agreed with the primary analysis, and that the 28-day mortality did not suffer from reporting bias according to the funnel plots. Second, our findings may potentially be biased by the evolving quality of care for sepsis^32 62-65^ because we still included trials published back in 1980s and1990s.^39 40 42 45^ Nevertheless, the sensitivity analysis excluding trials published before 2012 still agreed with the primary analysis. Third, we may underestimate the effects of corticosteroids in some subgroup analysis with fewer available data due to inadequate statistical power (Table 3). Lastly, we excluded clinical trials without adequate published inclusion details necessary for the Sepsis-3 criteria; therefore, patients fulfilling the Sepsis-3 definitions in these studies may be excluded and therefore were underrepresented in this study.

## Conclusion

This meta-analysis suggested that intravenous corticosteroids compared to standard of care or placebo did not significantly reduce the 28-day mortality and were associated with an increased risk of hyperglycemia in patients with sepsis defined by the Sepsis-3 criteria. Further studies are warranted to understand the roles of disease severity and treatment timing on the efficacy of corticosteroid treatment in this population.

## Data Availability

All data produced in the present work are contained in the manuscript

## Footnotes

## Acknowledgements

None

## Author contributions

Dr. Wu had full access to all the data in the study and takes responsibility for the integrity of the data and accuracy of the data analysis.

*Concept and design*: Y. Wu, and S. Papatheodorou.

*Acquisition, analysis, or interpretation of data*: All authors.

*Drafting of the manuscript*: Y. Wu

*Critical revision of the manuscript for important intellectual content*: Y. Wu, C. Lin, R. Hamaya, F. Huang, S. Chen, and S. Papatheodorou.

*Statistical analysis:* Y. Wu, and C. Lin

*Administrative, technical, or material support:* All authors.

*Supervision:* S. Papatheodorou

All authors have read and approved the content for submission.

## Patient consent

Not required.

## Data sharing statement

Extracted data are available by request to the corresponding author.

