## Supplementary Materials for "Intravenous corticosteroid treatment in adult patients with sepsis defined by the Sepsis-3 criteria: a systematic review and meta-analysis"

Wu Y, Lin C, Hamaya R, et al.

eTable 1. Search Strategy

eTable 2. Detailed Judgements on Trials with Some Concerns or High Risk of Bias

eTable 3. Justification of Fulfilling the Sepsis-3 Definition for Septic Shock

eFigure 1. Risk of Bias per Domain per Study

eFigure 2. Weighted Risk of Bias for the 28-day Mortality (Primary outcome)

eFigure 3. Funnel Plot with Harbord test for 28-day Mortality

eFigure 4. Forest Plot for the Sensitivity Analysis (Excluding trials with total participants <100)

eFigure 5. Forest Plot for the Sensitivity Analysis (Excluding trials with events < 10 in one of the treatment group)

eFigure 6. Forest Plot for the Sensitivity Analysis (Only overall low risk of bias)

eFigure 7. Forest Plot for the Sensitivity Analysis (Excluding not double-blind studies)

eFigure 8. Forest Plot for the Sensitivity Analysis (Excluding studies published before 2012)

eFigure 9. Forest Plot for the Sensitivity Analysis (Using random-effects model with DerSimonian-Laird method)

eFigure 10. Forest Plot for the Sensitivity Analysis (Fix-effects model)

eFigure 11. Forest Plot for Length of ICU Stay

eFigure 12. Forest Plot for Length of Hospital Stay

eFigure 13. Forest Plot for SOFA Score at Day 7

eFigure 14. Forest Plot for 90-day Mortality

eFigure 15. Forest Plot for In-hospital Mortality

eFigure 16. Forest Plot for ICU Mortality

eFigure 17. Forest Plot for Hyperglycemia

eFigure 18. Forest Plot for Gastrointestinal Bleeding

eFigure 19. Forest Plot for Superinfection

eFigure 20. Forest Plot for Any Adverse Event

eFigure 21. Funnel Plot with Egger’s test for Length of ICU Stay

eFigure 22. Funnel Plot with Egger’s test for Length of Hospital Stay

eFigure 23. Funnel Plot for SOFA Score at Day 7

eFigure 24. Funnel Plot for 90-day Mortality

eFigure 25. Funnel Plot with Harbord’s Test for In-hospital Mortality

eFigure 26. Funnel Plot with Harbord’s Test for ICU Mortality

eFigure 27. Funnel Plot with Harbord’s Test for Hyperglycemia

eFigure 28. Funnel Plot with Harbord’s Test for Gastrointestinal Bleeding

eFigure 29. Funnel Plot with Harbord’s Test for Superinfection

eFigure 30. Funnel Plot for Any Adverse Events

eFigure 31. Subgroup Analysis: Stratified by Treatment Duration ($\leq$7 days)

eFigure 32. Subgroup Analysis: Stratified by Treatment Duration ($>$7 days)

eFigure 33. Subgroup Analysis: Stratified by Treatment Duration ($\leq$7 days vs $>$7 days)

eFigure 34. Subgroup Analysis: Stratified by the Hydrocortisone Equivalent Dose ($\leq$200mg per day)

eFigure 35. Subgroup Analysis: Stratified by the Hydrocortisone Equivalent Dose ($>$200mg per day)

eFigure 36. Subgroup Analysis: Stratified by the Hydrocortisone Equivalent Dose ($\leq$200mg vs >200mg per day)

eFigure 37. Subgroup Analysis: Stratified by the Type of Corticosteroid (hydrocortisone)

eFigure 38. Subgroup Analysis: Stratified by the Type of Corticosteroid (methylprednisolone)

eFigure 39. Subgroup Analysis: Stratified by the Type of Corticosteroid (hydrocortisone plus fludrocortisone)

eFigure 40. Subgroup Analysis: Stratified by the Type of Corticosteroid (dexamethasone)

eFigure 41. Subgroup Analysis: Stratified by the Type of Corticosteroid (hydrocortisone vs methylprednisolone vs hydrocortisone plus fludrocortisone vs dexamethasone)

**eTable 1. Search Strategy**

| **PubMed (July 12th, 2019)** | | |
| --- | --- | --- |
| #1 | "Steroids"[Mesh] OR "Adrenal Cortex Hormones"[Mesh] OR steroid*[tiab] OR glucocorticoid[tiab] OR Corticoid*[tiab] OR Adrenal Cortex Hormone*[tiab] OR cortisol[tiab] OR cortison*[tiab] OR hydrocortison*[tiab] OR corticosteroid*[tiab] OR prednisolon*[tiab] OR methylprednisolon*[tiab] OR prednison*[tiab] OR dexamethason*[tiab] OR triamcinolon*[tiab] OR Betamethason*[tiab] OR Fludrocortison*[tiab] | 1121644 |
| #2 | "Systemic Inflammatory Response Syndrome"[Mesh] OR "Endocarditis, Bacterial"[Mesh] OR "pneumonia"[Mesh] OR "Respiratory Distress Syndrome, Adult"[Mesh] OR "Urinary Tract Infections"[Mesh] OR "Suppuration"[Mesh] OR "Cross Infection"[Mesh] OR "Community-Acquired Infections"[Mesh] OR Systemic Inflammatory Response Syndrome[tiab] OR SIRS[tiab] OR seps*[tiab] OR septic*[tiab] OR SOFA[tiab] OR Sequential Organ Failure Assessment[tiab] OR pneumonia[tiab] OR CAP[tiab] OR Respiratory Distress Syndrome[tiab] OR ARDS[tiab] | 582916 |
| #3 | "randomized controlled trial"[pt] OR "controlled clinical trial"[pt] OR randomized[tiab] OR placebo[tiab] OR "drug therapy"[sh] OR randomly[tiab] OR trial[tiab] OR groups[tiab] | 4549989 |
| #4 | ("Editorial"[pt] OR "Letter"[pt] "Comment"[pt] OR "case reports"[pt] OR "Review"[pt]) | 5007098 |
| #5 | ("Animals"[Mesh] NOT "Humans"[Mesh]) | 4604516 |
| #6 | #1 AND #2 AND #3 NOT #4 NOT #5 | 5846 |
| **PubMed (June 28th, 2020)** | | |
| #1 | "Steroids"[Mesh] OR "Adrenal Cortex Hormones"[Mesh] OR steroid*[tiab] OR glucocorticoid[tiab] OR Corticoid*[tiab] OR Adrenal Cortex Hormone*[tiab] OR cortisol[tiab] OR cortison*[tiab] OR hydrocortison*[tiab] OR corticosteroid*[tiab] OR prednisolon*[tiab] OR methylprednisolon*[tiab] OR prednison*[tiab] OR dexamethason*[tiab] OR triamcinolon*[tiab] OR Betamethason*[tiab] OR Fludrocortison*[tiab] | 1153424 |
| #2 | "Systemic Inflammatory Response Syndrome"[Mesh] OR "Endocarditis, Bacterial"[Mesh] OR "pneumonia"[Mesh] OR "Respiratory Distress Syndrome, Adult"[Mesh] OR "Urinary Tract Infections"[Mesh] OR "Suppuration"[Mesh] OR "Cross Infection"[Mesh] OR "Community-Acquired Infections"[Mesh] OR Systemic Inflammatory Response Syndrome[tiab] OR SIRS[tiab] OR seps*[tiab] OR septic*[tiab] OR SOFA[tiab] OR Sequential Organ Failure Assessment[tiab] OR pneumonia[tiab] OR CAP[tiab] OR Respiratory Distress Syndrome[tiab] OR ARDS[tiab] | 618630 |
| #3 | "randomized controlled trial"[pt] OR "controlled clinical trial"[pt] OR randomized[tiab] OR placebo[tiab] OR "drug therapy"[sh] OR randomly[tiab] OR trial[tiab] OR groups[tiab] | 4794425 |
| #4 | ("Editorial"[pt] OR "Letter"[pt] "Comment"[pt] OR "case reports"[pt] OR "Review"[pt]) | 5238606 |
| #5 | ("Animals"[Mesh] NOT "Humans"[Mesh]) | 4712175 |
| #6 | #1 AND #2 AND #3 NOT #4 NOT #5 | 6234 |
| **Embase (July 12th, 2019)** | | |
| #1 | Steroid/exp OR steroid*:ab,ti OR glucocorticoid:ab,ti OR Corticoid*:ab,ti OR ‘Adrenal Cortex Hormone*’:ab,ti OR cortisol:ab,ti OR cortison*:ab,ti OR hydrocortison*:ab,ti OR corticosteroid*:ab,ti OR prednisolon*:ab,ti OR methylprednisolon*:ab,ti OR prednison*:ab,ti OR dexamethason*:ab,ti OR triamcinolon*:ab,ti OR Betamethason*:ab,ti OR Fludrocortison*:ab,ti | 1405828 |
| #2 | ‘Systemic Inflammatory Response Syndrome’/exp OR ‘Systemic Inflammatory Response Syndrome’:ab,ti OR SIRS:ab,ti OR seps*:ab,ti OR septic*:ab,ti OR ‘SOFA’:ab,ti OR ‘Sequential Organ Failure Assessment‘:ab,ti OR ‘bloodstream infection’/exp OR ‘community acquired infection’/exp OR ‘lower respiratory tract infection’/exp OR ‘urinary tract infection’/exp OR ‘bacterial endocarditis’/exp OR ‘cross infection’/exp OR ‘suppuration’/exp OR ’adult respiratory distress syndrome’/exp OR ‘pneumonia’/exp | 692692 |
| #3 | 'randomized controlled trial'/exp OR 'controlled clinical trial'/exp OR randomized:ti,ab OR placebo:ti,ab OR randomly:ti,ab OR trial:ti,ab OR groups:ti,ab | 3286027 |
| #4 | ([conference abstract]/lim OR [conference review]/lim OR [editorial]/lim OR [letter]/lim OR [review]/lim OR [short survey]/lim) | 7200515 |
| #5 | ([animals]/lim NOT [humans]/lim) | 4077256 |
| #6 | #1 AND #2 AND #3 NOT 4 NOT #5 | 8023 |
| **Embase (June 28th, 2020)** | | |
| #1 | Steroid/exp OR steroid*:ab,ti OR glucocorticoid:ab,ti OR Corticoid*:ab,ti OR ‘Adrenal Cortex Hormone*’:ab,ti OR cortisol:ab,ti OR cortison*:ab,ti OR hydrocortison*:ab,ti OR corticosteroid*:ab,ti OR prednisolon*:ab,ti OR methylprednisolon*:ab,ti OR prednison*:ab,ti OR dexamethason*:ab,ti OR triamcinolon*:ab,ti OR Betamethason*:ab,ti OR Fludrocortison*:ab,ti | 1828263 |
| #2 | ‘Systemic Inflammatory Response Syndrome’/exp OR ‘Systemic Inflammatory Response Syndrome’:ab,ti OR SIRS:ab,ti OR seps*:ab,ti OR septic*:ab,ti OR ‘SOFA’:ab,ti OR ‘Sequential Organ Failure Assessment‘:ab,ti OR ‘bloodstream infection’/exp OR ‘community acquired infection’/exp OR ‘lower respiratory tract infection’/exp OR ‘urinary tract infection’/exp OR ‘bacterial endocarditis’/exp OR ‘cross infection’/exp OR ‘suppuration’/exp OR ’adult respiratory distress syndrome’/exp OR ‘pneumonia’/exp | 977997 |
| #3 | 'randomized controlled trial'/exp OR 'controlled clinical trial'/exp OR randomized:ti,ab OR placebo:ti,ab OR randomly:ti,ab OR trial:ti,ab OR groups:ti,ab | 4411581 |
| #4 | ([conference abstract]/lim OR [conference review]/lim OR [editorial]/lim OR [letter]/lim OR [review]/lim OR [short survey]/lim) | 8854425 |
| #5 | ([animals]/lim NOT [humans]/lim) | 6045512 |
| #6 | #1 AND #2 AND #3 NOT 4 NOT #5 | 9577 |
| **Cochrane Central Register of Controlled Trials (July 12th, 2019)** | | |
| #1 | (steroid* OR glucocorticoid OR Corticoid* OR Adrenal Cortex Hormone* OR cortisol OR cortison* OR hydrocortison* OR corticosteroid* OR prednisolon* OR methylprednisolon* OR prednison* OR dexamethason* OR triamcinolon* OR Betamethason* OR Fludrocortison*) | 81608 |
| #2 | (Systemic Inflammatory Response Syndrome OR SIRS OR seps* OR septic* OR SOFA OR Sequential Organ Failure Assessment OR bloodstream infection OR community acquired infection OR pneumonia OR CAP OR lower respiratory tract infection OR urinary tract infection OR bacterial endocarditis OR cross infection OR suppuration OR adult respiratory distress syndrome OR acute respiratory distress syndrome OR ARDS) | 48880 |
| #3 | #1 AND #2 AND only trials | 3613 |
| **Cochrane Central Register of Controlled Trials (June 28th, 2020)** | | |
| #1 | (steroid* OR glucocorticoid OR Corticoid* OR Adrenal Cortex Hormone* OR cortisol OR cortison* OR hydrocortison* OR corticosteroid* OR prednisolon* OR methylprednisolon* OR prednison* OR dexamethason* OR triamcinolon* OR Betamethason* OR Fludrocortison*) | 82975 |
| #2 | (Systemic Inflammatory Response Syndrome OR SIRS OR seps* OR septic* OR SOFA OR Sequential Organ Failure Assessment OR bloodstream infection OR community acquired infection OR pneumonia OR CAP OR lower respiratory tract infection OR urinary tract infection OR bacterial endocarditis OR cross infection OR suppuration OR adult respiratory distress syndrome OR acute respiratory distress syndrome OR ARDS) | 52724 |
| #3 | #1 AND #2 AND only trials | 4142 |
| **Web of Science (July 12th, 2019)** | | |
| #1 | (steroid* OR glucocorticoid OR Corticoid* OR Adrenal Cortex Hormone* OR cortisol OR cortison* OR hydrocortison* OR corticosteroid* OR prednisolon* OR methylprednisolon* OR prednison* OR dexamethason* OR triamcinolon* OR Betamethason* OR Fludrocortison*) | 534885 |
| #2 | (Systemic Inflammatory Response Syndrome OR SIRS OR seps* OR septic* OR SOFA OR Sequential Organ Failure Assessment OR bloodstream infection OR community acquired infection OR lower respiratory tract infection OR urinary tract infection OR bacterial endocarditis OR cross infection OR suppuration OR adult respiratory distress syndrome) | 343713 |
| #3 | (randomized controlled trial OR controlled clinical trial OR randomized OR  placebo OR randomly OR trial OR groups) | 5994963 |
| #4 | SU=Veterinary Sciences | 654063 |
| #5 | SU=Zoology | 585781 |
| #6 | #1 AND #2 AND #3 NOT #4 NOT #5 | 3602 |
| **Web of Science (June 28th, 2020)** | | |
| #1 | (steroid* OR glucocorticoid OR Corticoid* OR Adrenal Cortex Hormone* OR cortisol OR cortison* OR hydrocortison* OR corticosteroid* OR prednisolon* OR methylprednisolon* OR prednison* OR dexamethason* OR triamcinolon* OR Betamethason* OR Fludrocortison*) | 15588 |
| #2 | (Systemic Inflammatory Response Syndrome OR SIRS OR seps* OR septic* OR SOFA OR Sequential Organ Failure Assessment OR bloodstream infection OR community acquired infection OR lower respiratory tract infection OR urinary tract infection OR bacterial endocarditis OR cross infection OR suppuration OR adult respiratory distress syndrome) | 26383 |
| #3 | (randomized controlled trial OR controlled clinical trial OR randomized OR  placebo OR randomly OR trial OR groups) | 663794 |
| #4 | SU=Veterinary Sciences | 15933 |
| #5 | SU=Zoology | 17613 |
| #6 | #1 AND #2 AND #3 NOT #4 NOT #5 AND Document type: trials | 1577 |
| **ClinicalTrials.gov (July 12th, 2019)** | | |
| #1 | (steroid OR glucocorticoid OR Corticoid OR Adrenal Cortex Hormone OR cortisol OR cortisone OR hydrocortisone OR corticosteroide OR prednisolone OR methylprednisolone OR prednisone OR dexamethasone OR triamcinolone OR Betamethasone OR Fludrocortisone) | -- |
| #2 | (Systemic Inflammatory Response Syndrome OR SIRS OR sepsis OR septic OR SOFA OR Sequential Organ Failure Assessment OR bloodstream infection OR community acquired infection OR lower respiratory tract infection OR urinary tract infection) | -- |
| #3 | (bacterial endocarditis OR cross infection OR suppuration OR adult respiratory distress syndrome) | -- |
| #4 | #1 AND #2 | 165 |
| #5 | #1 AND #3 | 71 |
| #6 | #4 + #5 | 236 |
| **ClinicalTrials.gov (June 28th, 2020)** | | |
| #1 | (steroid OR glucocorticoid OR Corticoid OR Adrenal Cortex Hormone OR cortisol OR cortisone OR hydrocortisone OR corticosteroide OR prednisolone OR methylprednisolone OR prednisone OR dexamethasone OR triamcinolone OR Betamethasone OR Fludrocortisone) | -- |
| #2 | (Systemic Inflammatory Response Syndrome OR SIRS OR sepsis OR septic OR SOFA OR Sequential Organ Failure Assessment OR bloodstream infection OR community acquired infection OR lower respiratory tract infection OR urinary tract infection) | -- |
| #3 | (bacterial endocarditis OR cross infection OR suppuration OR adult respiratory distress syndrome) | -- |
| #4 | #1 AND #2 | 186 |
| #5 | #1 AND #3 | 90 |
| #6 | #4 + #5 | 276 |
| **International Clinical Trials Registry Platform (July 12th, 2019)^a^** | | |
| #1 | For the title:  (steroid* OR glucocorticoid OR Corticoid* OR Adrenal Cortex Hormone* OR cortisol OR cortison* OR hydrocortison* OR corticosteroid* OR prednisolon* OR methylprednisolon* OR prednison* OR dexamethason* OR triamcinolon* OR Betamethason* OR Fludrocortison*) AND (Systemic Inflammatory Response Syndrome OR SIRS OR seps* OR septic* OR SOFA OR Sequential Organ Failure Assessment OR bloodstream infection) | 73 |
| #2 | For the title:  (steroid* OR glucocorticoid OR Corticoid* OR Adrenal Cortex Hormone* OR cortisol OR cortison* OR hydrocortison* OR corticosteroid* OR prednisolon* OR methylprednisolon* OR prednison* OR dexamethason* OR triamcinolon* OR Betamethason* OR Fludrocortison*) AND (community acquired infection OR lower respiratory tract infection OR urinary tract infection OR bacterial endocarditis OR cross infection OR suppuration OR adult respiratory distress syndrome) | 9 |
| #3 | For the intervention:  (steroid* OR glucocorticoid OR Corticoid* OR Adrenal Cortex Hormone* OR cortisol OR cortison* OR hydrocortison* OR corticosteroid* OR prednisolon* OR methylprednisolon* OR prednison* OR dexamethason* OR triamcinolon* OR Betamethason* OR Fludrocortison*) | -- |
| #4 | For the condition:  (Systemic Inflammatory Response Syndrome OR SIRS OR seps* OR septic* OR SOFA OR Sequential Organ Failure Assessment OR bloodstream infection) | -- |
| #5 | #3 AND #4 | 79 |
| #6 | For the condition:  (community acquired infection OR lower respiratory tract infection OR urinary tract infection OR bacterial endocarditis OR adult respiratory distress syndrome) |  |
| #7 | #3 AND #6 | 797 |
| #8 | #1 + #2 + #5 + #7 | 958 |
| ^a^Advanced Search of the International Clinical Trials Registry Platform was unavailable on June 28^th^, 2020, so we used the results obtained on July 12^th^, 2019 instead. | | |

**eTable 2. Detailed Judgements on Trials with Some Concerns or High Risk of Bias**

| **Study** | **Domain:** | **Overall** |
| --- | --- | --- |
| Sabry 2011^1^ | D1, No information about the concealment available;  D1, No information on how the randomization was done; | Some concerns |
| Cicarelli 2007^2^ | D1, Based on Table 1, days of vasopressor use was significantly different;  D5, No trial registration was found. | Some concerns |
| Lv 2017^3^ | D1, Based on Table 1, most of the baseline characteristics are similar. However, the initial APACHE II score and SOFA score were significantly different between the two groups, which may influence results;  D1, No information about the concealment available;  D5, Based on the trial registration (NCT02580240), the pre-specified secondary outcome, death from any cause at 90 days, was not reported in the Journal article. Also, reported secondary outcomes such as the reversal of shock, in-hospital mortality, duration of hospital and ICU stay were not mentioned in the trial registration. | Some concerns |
| Fernández-Serrano 2011^4^ | D1, No information about the concealment available;  D3, 11 out of 56 individuals were not analyzed after randomization;  D5, This tiral was registered before the analysis (ISRCTN22426306). However, in the registration, early or late mortality were not defined. | High |
| CSG 1963^5^ | D1, no information on how the randomization was done;  D1, no distribution of baseline characteristics for the two groups was provided;  D5, No trial registration was available. | High |
| Confalonieri 2005^6^ | D1, Based on Table 1, some baseline characteristics were not balanced, which may influence results;  D5, No trial registration was available. | Some concerns |
| Annane 2018^7^ | D5, Some outcomes listed in the protocol or registration were not reported in the article, such as neurological sequels at 90 days and 180 days. | Some concerns |
| Oppert 2005^8^ | D1, According to Table 1, some baseline characteristics seem to be not balanced, such as age, site of infection, organ dysfunction, and cortisol;  D1, no information on how the randomization was done;  D3, 7 participants (7/48) were excluded from the analyses;  D5, No trial registration was available. | High |
| Arabi 2010^9^ | D5, Trial was terminated prematurely; | High |
| Bollaert 1998^10^ | D5, Trial was terminated prematurely; | High |
| Briegel 1999^11^ | D5, No trial registration was available. | Some concerns |
| Hu 2009^12^ | D1, Very few baseline characteristics were compared;  D1, No information about the concealment available;  D5, No trial registration was available. | High |
| Sprung 1984^13^ | D2, Not using a double-blinded setting: "University of Miami Research Committee would not allow the study to be performed in a double-blind manner"  D4, Not using a double-blinded setting;  D5, No trial registration was available. | High |
| Gordon 2014^14^ | D1, According to Table 1, some of the baseline characteristics seem not to be balanced, such as ethnicity, preexisting conditions, or organ failure.  D2, open-label;  D4, open-label. | High |
| Li 2016^15^ | D1, Very few baseline characteristics were reported and compared;  D2, No blinded information available;  D4, No blinded information available;  D5, No trial registration was available. | High |
| **Study** | **Domain:** | **Overall** |
| Chawla 1999^16^ | D1, No information about the concealment available;  D5, No trial registration was available. | Some concerns |
| Fan  2019^17^ | D1, only abstract, no data available for the baseline characteristics;  D2, single-blinded;  D3, no information about missing outcome data available;  D4, single-blinded;  D5, No trial registration was available. | High |
| Wani  2020^18^ | D2, open-label;  D4, open-label; | High |

D1, Bias arising from the randomization process;

D2, Bias due to deviations from intended interventions;

D3, Bias due to missing outcome data;

D4, Bias in measurement of the outcome;

D5, Bias in selection of the reported result;

**eTable 3. Justification of Fulfilling the Sepsis-3 Definition for Septic Shock**

| **Trial** | **Justification of Septic Shock^a^** | **Trial** | **Justification of Septic Shock^a^** |
| --- | --- | --- | --- |
| CSG  1963^5^ | No^b^ | Sprung  1984^13^ | Some subjects fulfilled^c^ |
| Bollaert  1998^10^ | Some subjects fulfilled^d^ | Chawla  1999^16,e^ | No^b^ |
| Briegel  1999^11^ | Some subjects fulfilled^f^ | Annane  2002^19^ | Some subjects fulfilled^g^ |
| Oppert  2005^8^ | No^b^ | Confalonieri  2005^6^ | No^h^ |
| Cicarelli  2007^2^ | No^b^ | Sprung  2008^20^ | Some subjects fulfilled^i^ |
| Hu  2009^12^ | No^b^ | Arabi  2010^9^ | Some subjects fulfilled^j^ |
| Sabry  2011^1^ | No^h^ | Fernández-Serrano  2011^4^ | No^h^ |
| Gordon  2014^14^ | Some subjects fulfilled^k^ | Gordon  2016^21^ | No^b^ |
| Tongyoo  2016^22^ | Some subjects fulfilled^l^ | Li  2016^15^ | No^b^ |
| Lv  2017^3^ | No^h^ | Annane  2018^7^ | Some subjects fulfilled^m^ |
| Doluee  2018^23^ | No^b^ | Venkatesh  2018^24^ | Some subjects fulfilled^n^ |
| Fan  2019^17,e^ | All subjects fulfilled | Wani  2020^18^ | Some subjects fulfilled^o^ |
| ^a^SOFA score$\geq$2 + vasopressor use + Serum lactate level > 2mmol/L;  ^b^No plasma lactate data available;  ^c^93% of patients used vasopressor, and the plasma lactate (mmol/L) in the three groups, mean (SD): 6.7 (2.0); 5.1 (1.1); 8.1(1.15);  ^d^Plasma lactate (mmol/L) in the two groups, median (range): 2.0 (0.9, 4.6); 1.9 (0.9, 8.0);  ^e^Abstract only  ^f^Plasma lactate (mmol/L) in the two groups, mean (SD): 3.8 (0.8); 3.4 (1.0);  ^g^Arterial lactate (mmol/L) of the two groups, mean (SD): 4.3 (4.3); 4.6 (4.4);  ^h^Not all patients used vasopressor and no plasma lactate data reported;  ^i^Over 80% of patients used vasopressor, and the plasma lactate (mmol/L) in the two groups, mean (SD): 3.9 (3.6); 4.1 (4.1);  ^j^All patients used vasopressor, and the plasma lactate (mmol/L) in the two groups, median (IQR): 2.8 (3.2); 3.0 (3.4);  ^k^Plasma lactate (mmol/L) in the two groups, median (range): 2.1 (1.4, 5.0); 1.8 (1.3, 2.7);  ^l^Over 76% of patients used vasopressor, and over 80% of patients with lactate >2 mmol/L;  ^m^Plasma lactate (mmol/L) of all patients, mean (SD): 4.36 (4.94);  ^n^Around 75% of patients had a plasma lactate concentration of 2 mmol/L;  ^o^76% and 92% of participants in the control and treatment group met the Sepsis-3 criteria for septic shock, respectively; | | | |

**eFigure 1. Risk of Bias per Domain per Study**

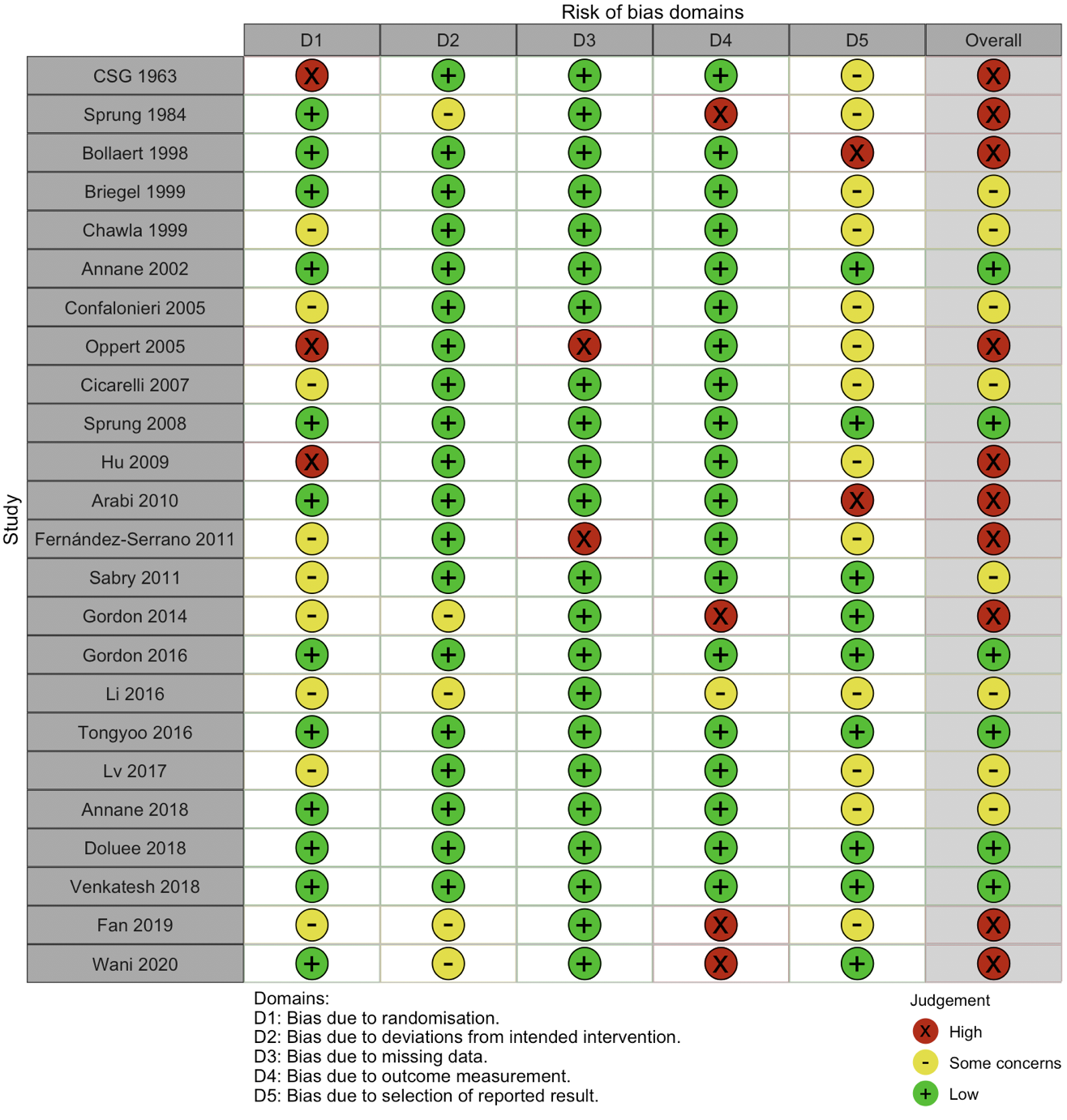

**eFigure 2. Weighted Risk of Bias for the 28-day Mortality (Primary outcome).**

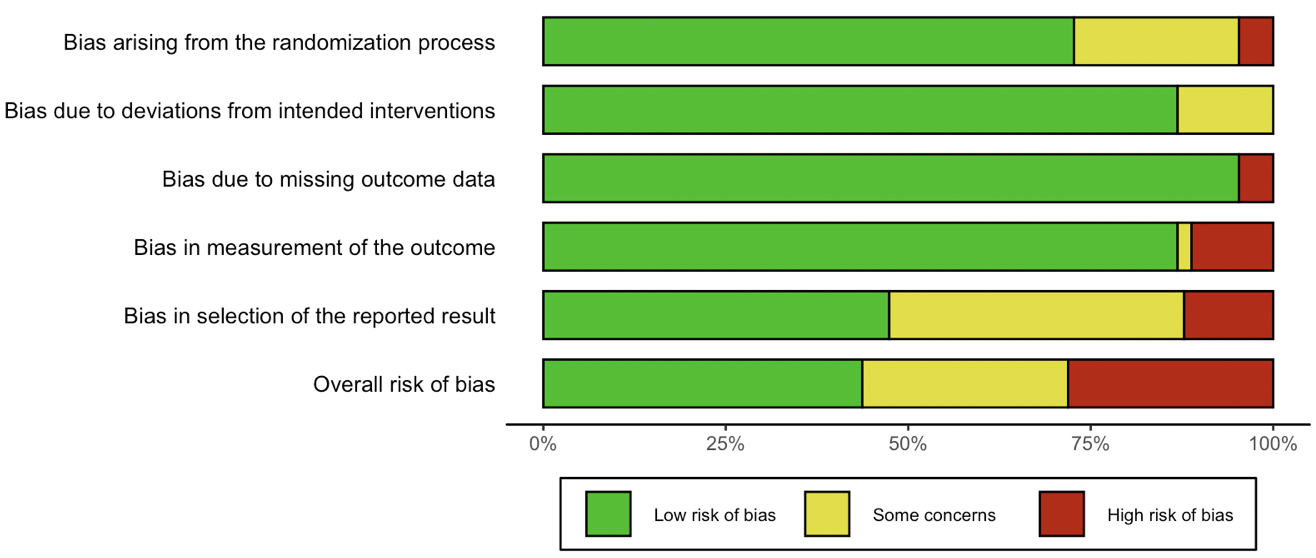

**eFigure 3. Funnel Plot with Harbord test for 28-day Mortality**

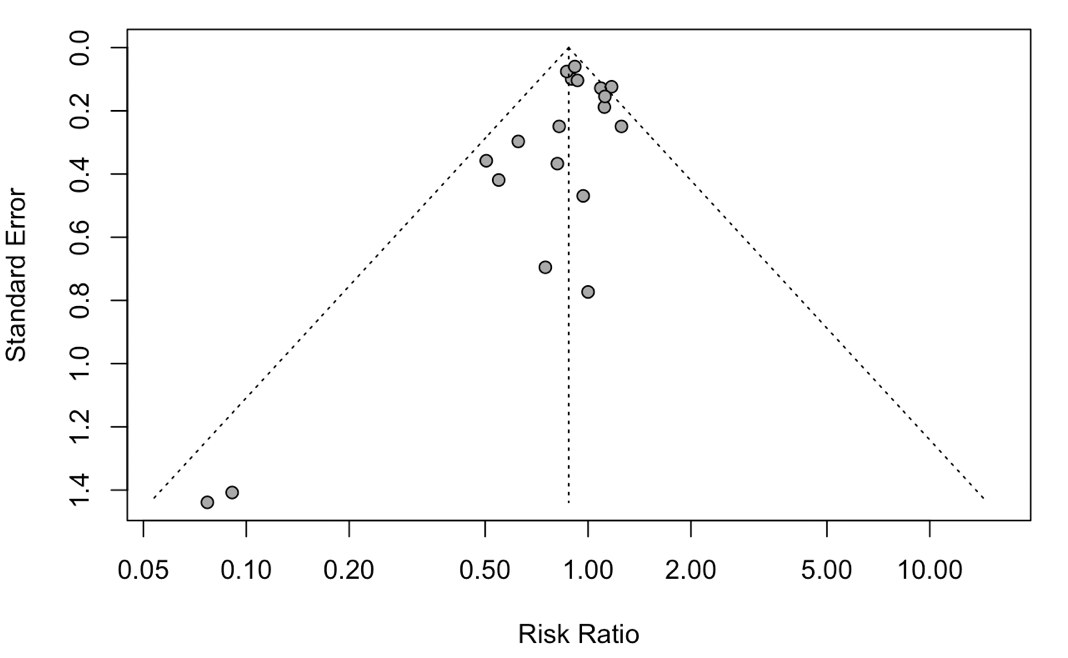

Harbord test: p = 0.11

**eFigure 4. Forest Plot for the Sensitivity Analysis (Excluding trials with total participants <100)**

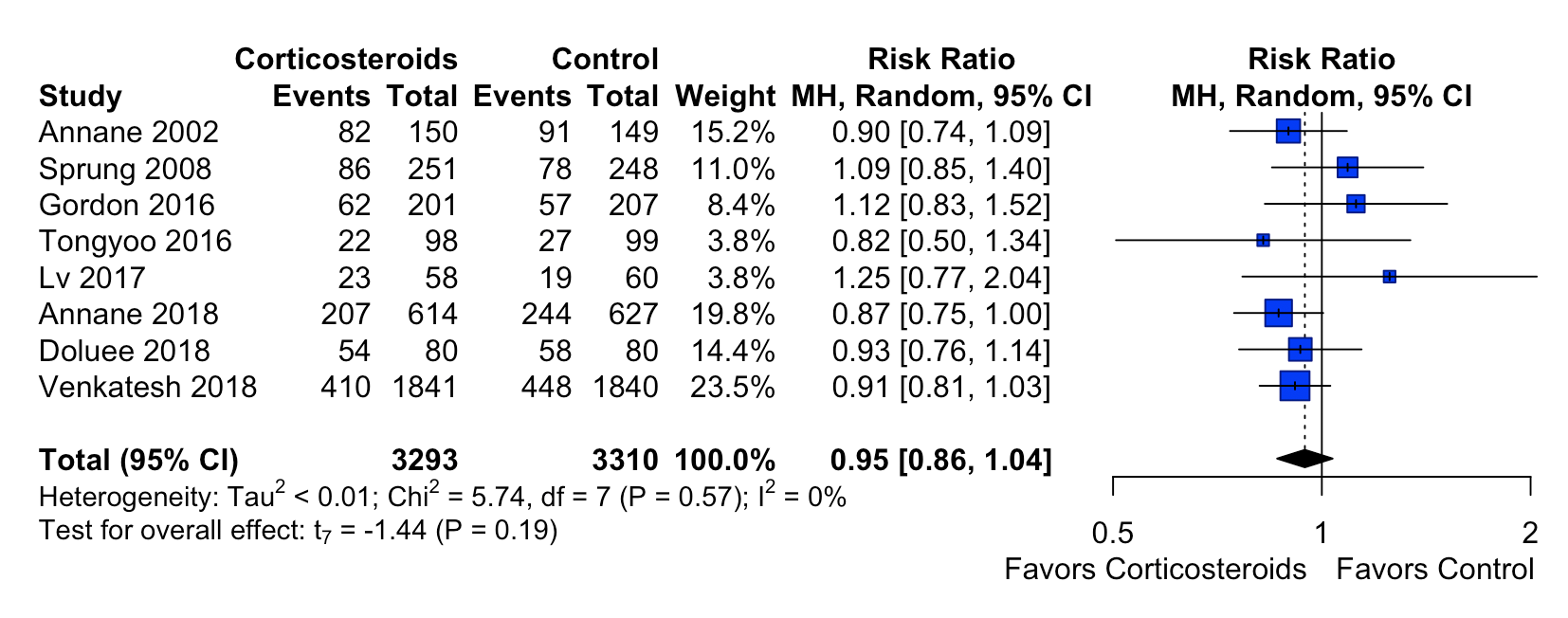

df, degrees of freedom; MH, Mantel-Haenszel method

**eFigure 5. Forest Plot for the Sensitivity Analysis (Excluding trials with events < 10 in one of the treatment group)**

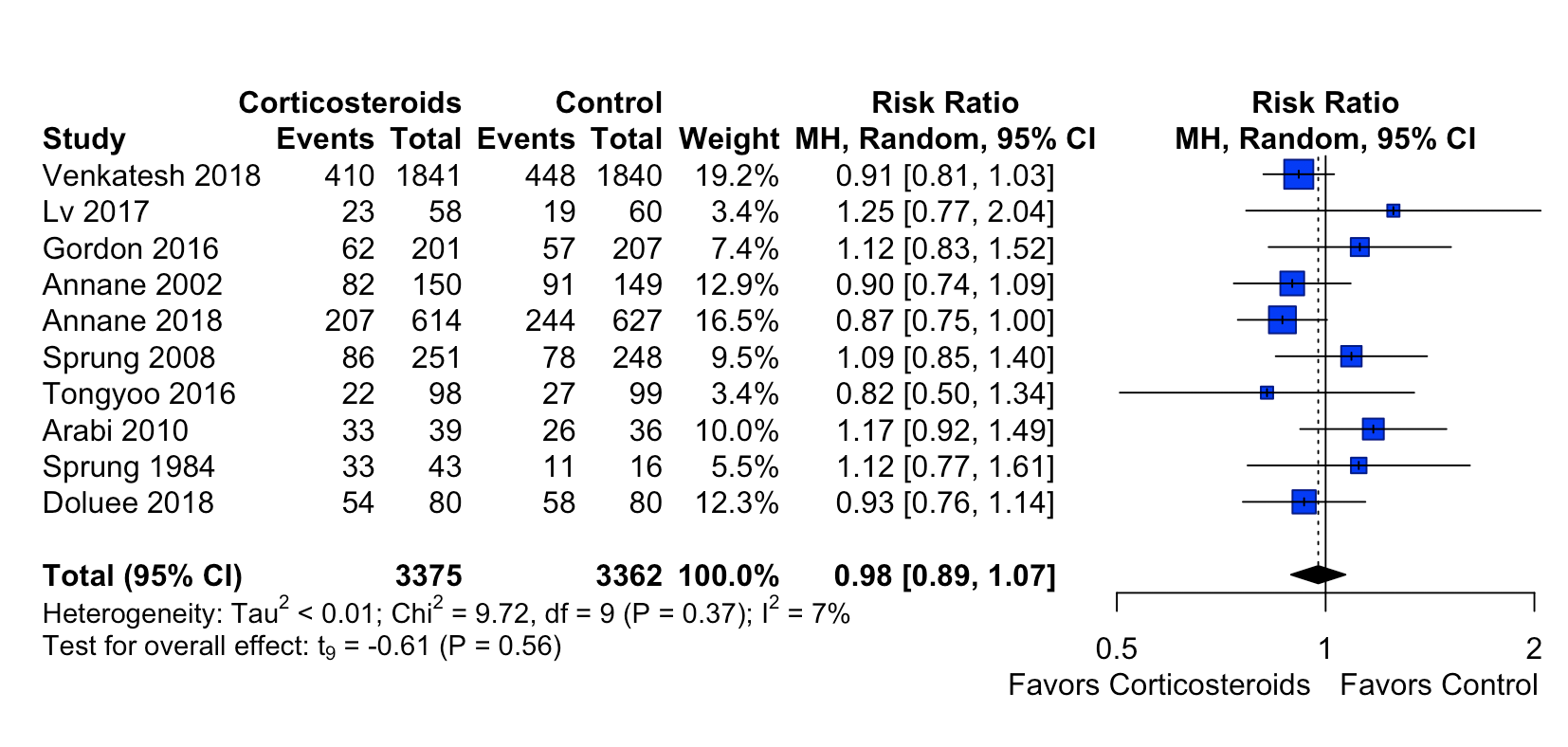

df, degrees of freedom; MH, Mantel-Haenszel method

**eFigure 6. Forest Plot for the Sensitivity Analysis (Only overall low risk of bias)**

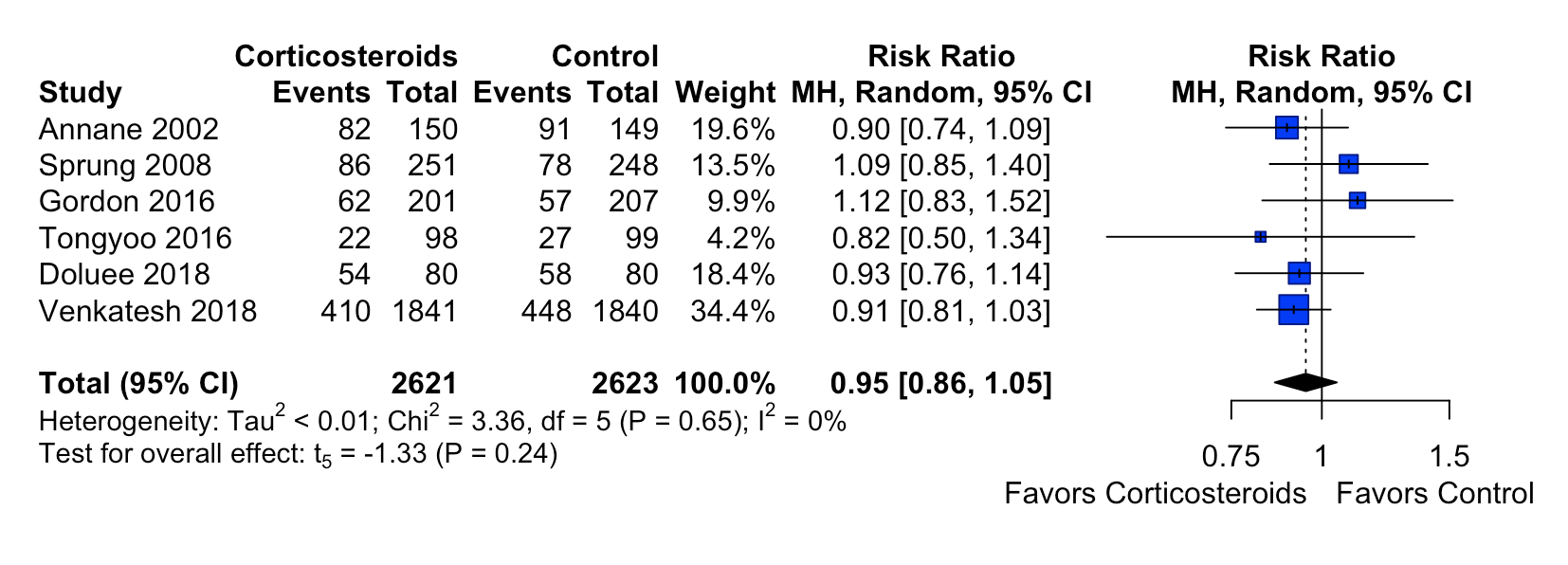

df, degrees of freedom; MH, Mantel-Haenszel method

**eFigure 7. Forest Plot for the Sensitivity Analysis (Excluding not double-blind studies)**

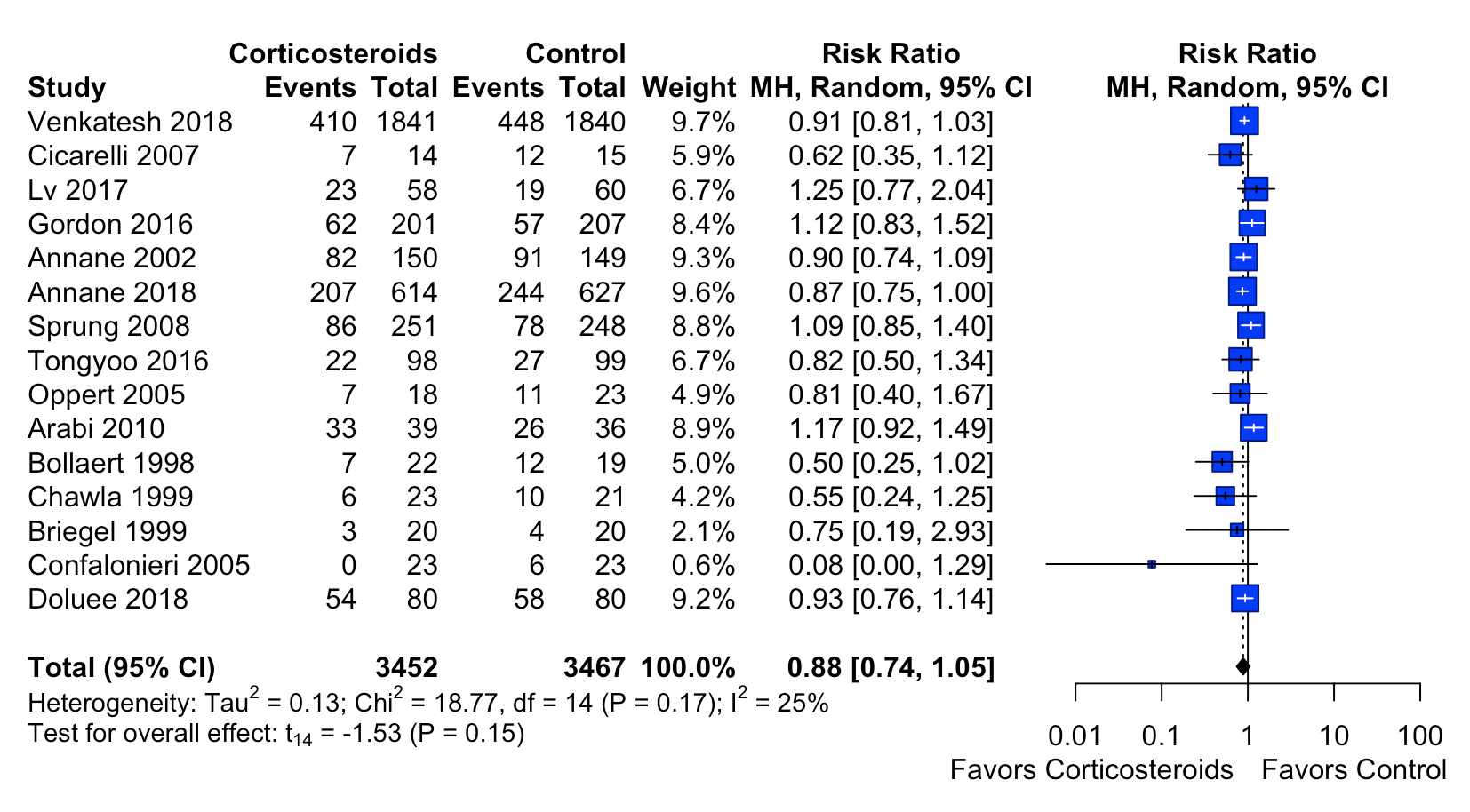

df, degrees of freedom; MH, Mantel-Haenszel method

**eFigure 8. Forest Plot for the Sensitivity Analysis (Excluding studies published before 2012)**

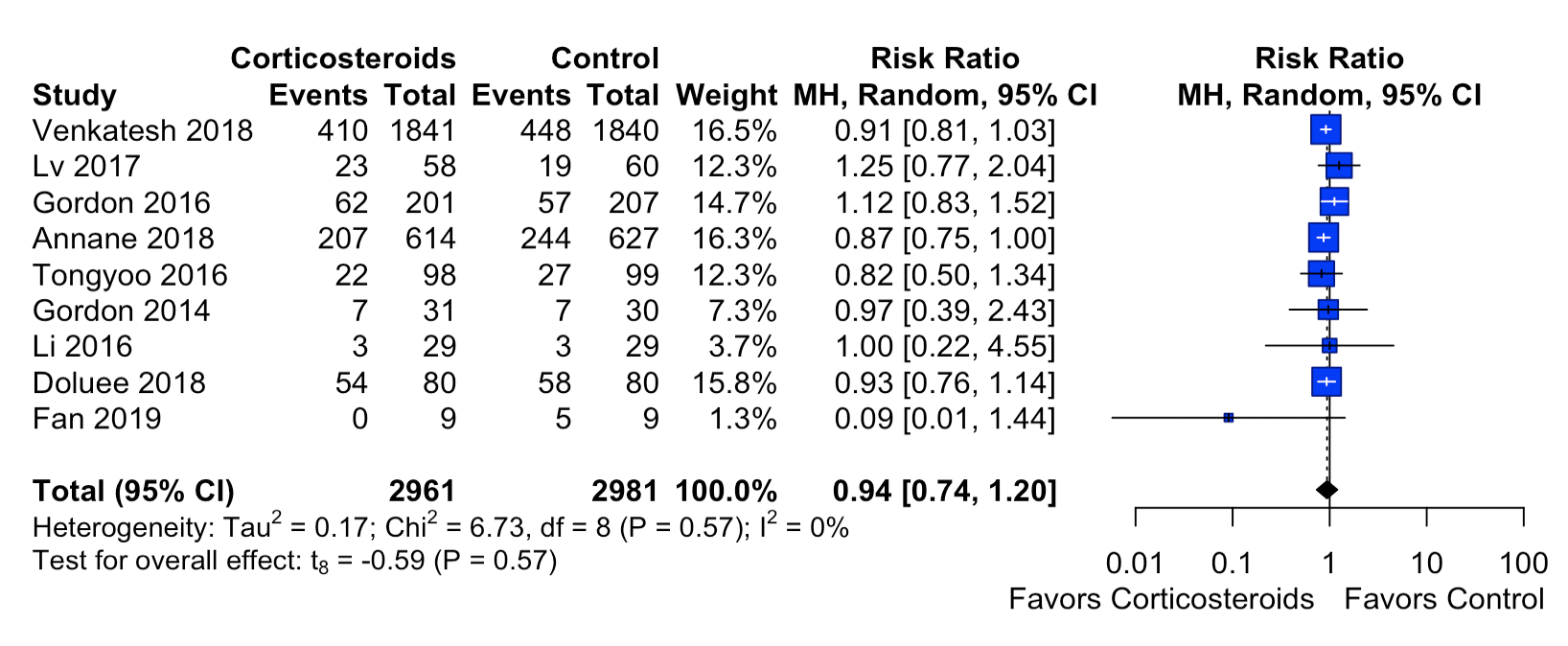

df, degrees of freedom; MH, Mantel-Haenszel method

**eFigure 9. Forest Plot for the Sensitivity Analysis (Using random-effects model with DerSimonian-Laird method)**

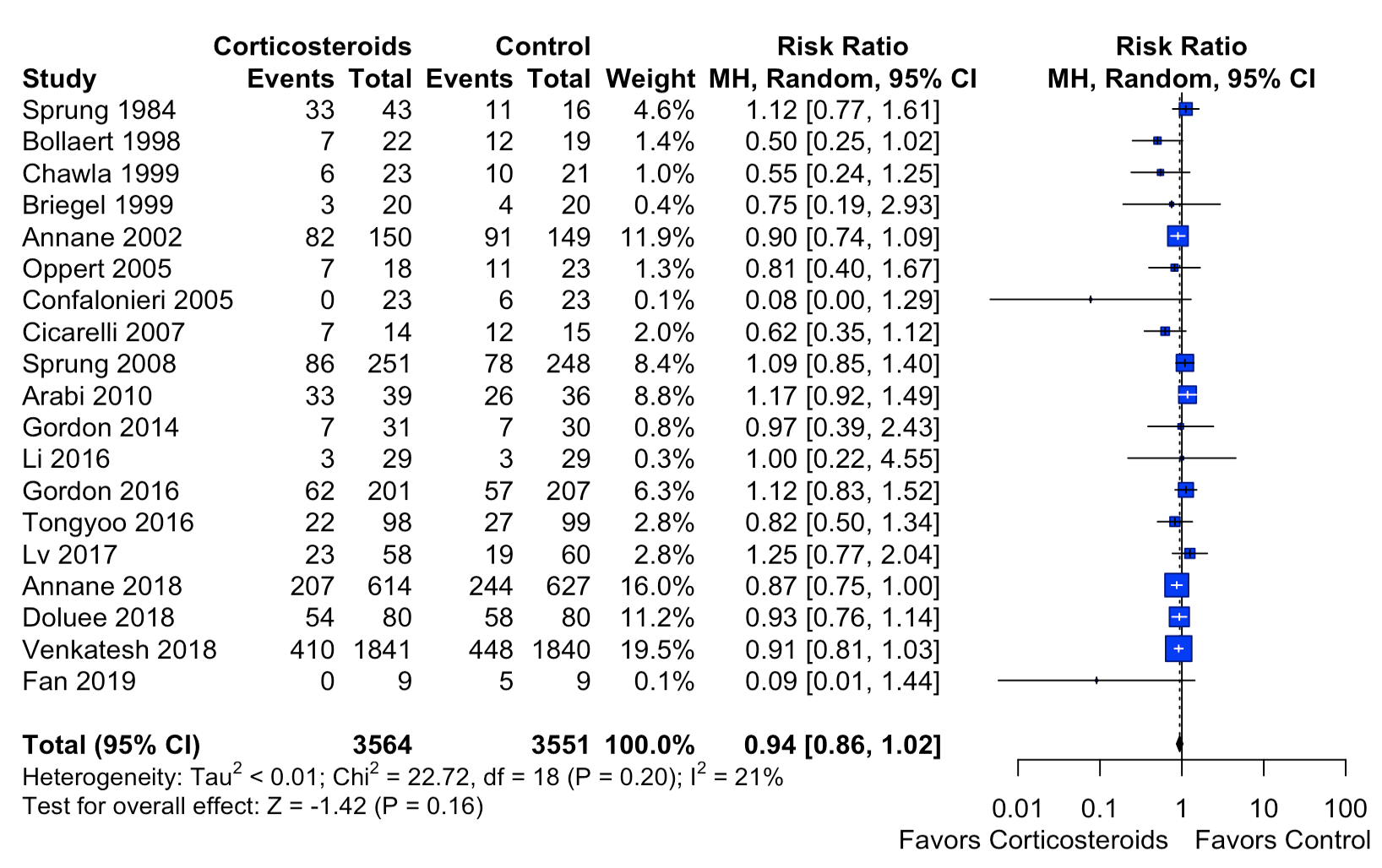

df, degrees of freedom; MH, Mantel-Haenszel method

### eFigure 10. Forest Plot for the Sensitivity Analysis (Fix-effects model)

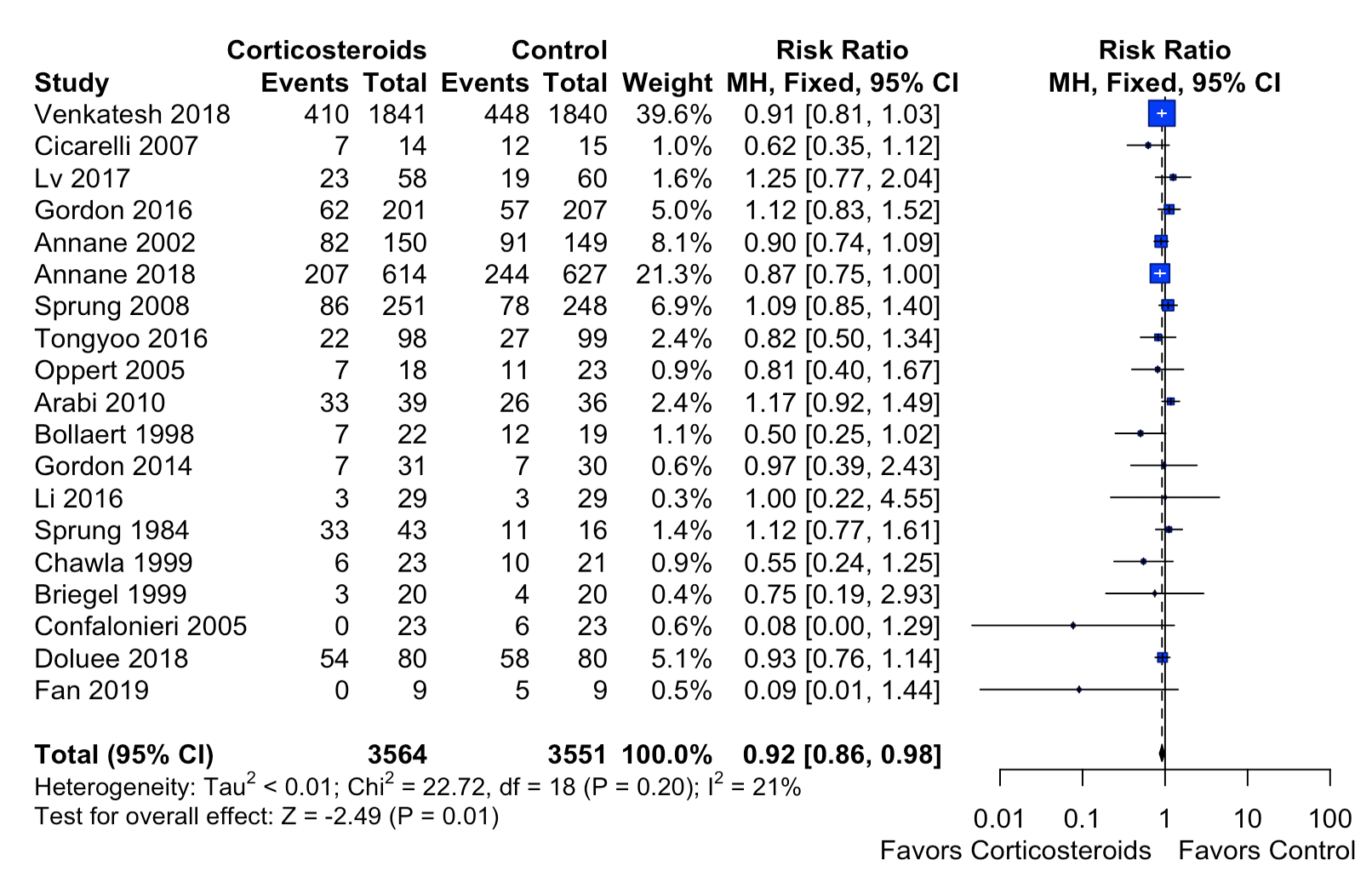

df, degrees of freedom; MH, Mantel-Haenszel method

**eFigure 11. Forest Plot for Length of ICU Stay**

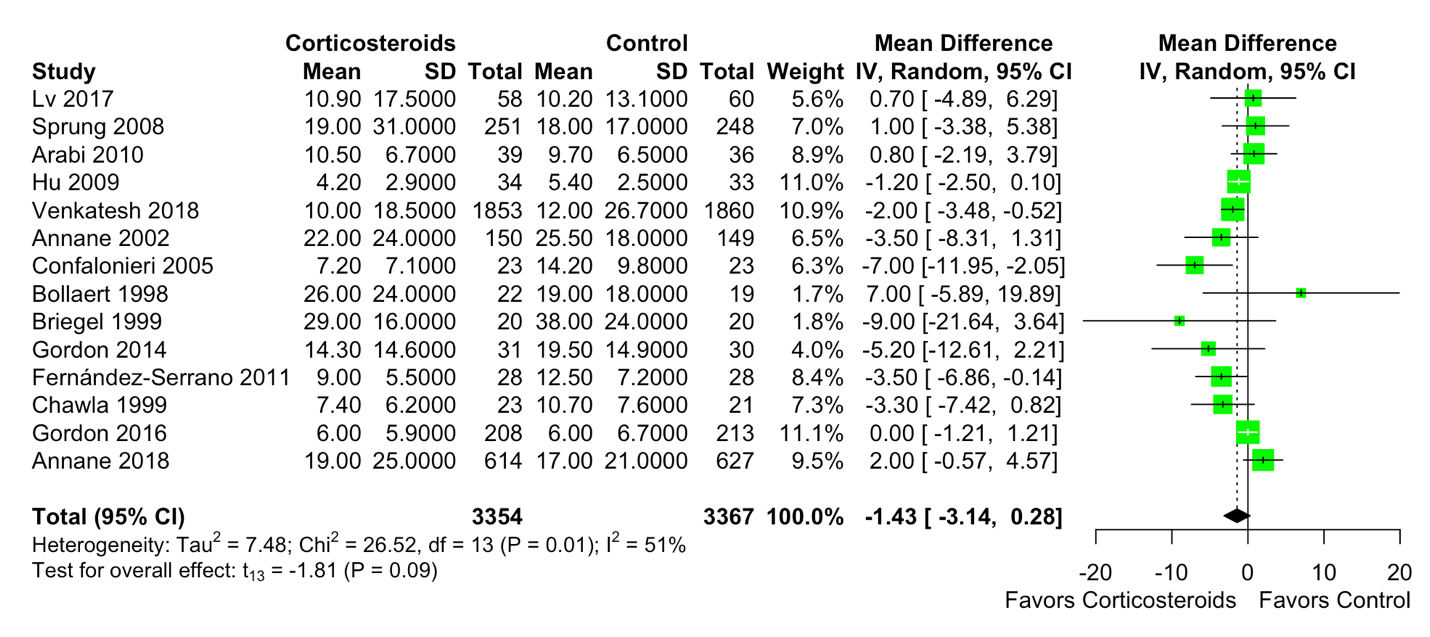

df, degrees of freedom; IV, inverse variance method

**eFigure 12. Forest Plot for Length of Hospital Stay**

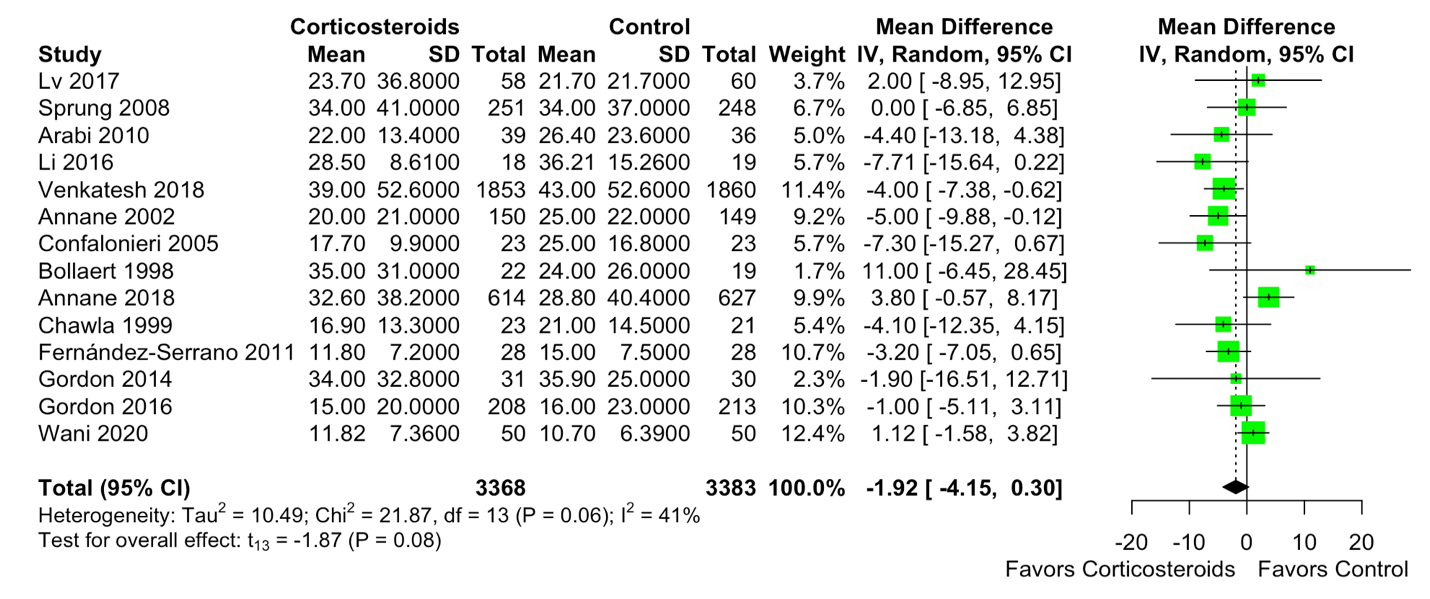

df, degrees of freedom; IV, inverse variance method

**eFigure 13. Forest Plot for SOFA Score at Day 7**

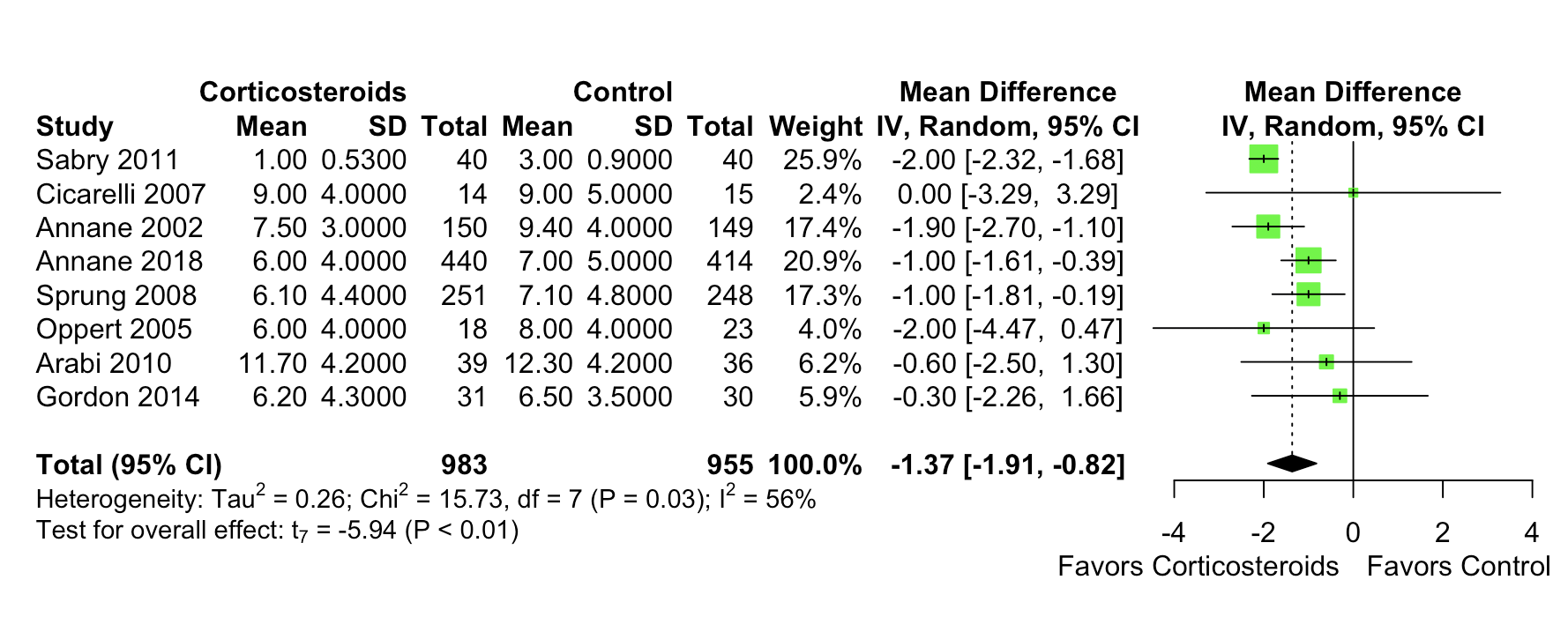

df, degrees of freedom; IV, inverse variance method

**eFigure 14. Forest Plot for 90-day Mortality**

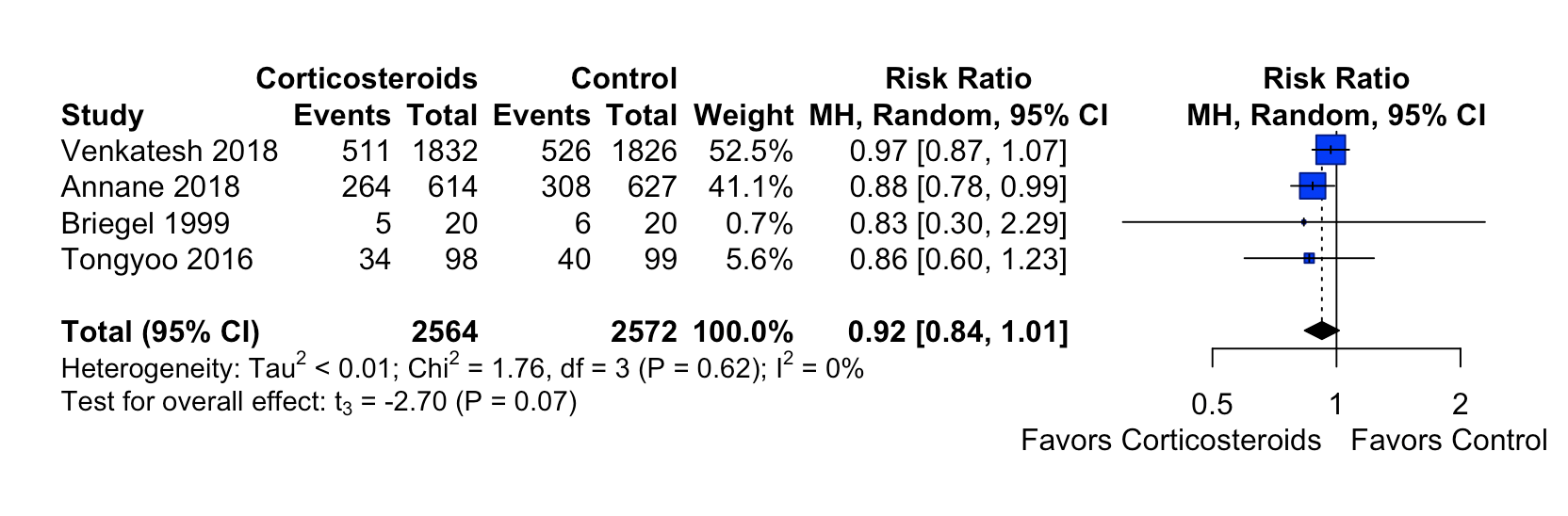

df, degrees of freedom; MH, Mantel-Haenszel method

**eFigure 15. Forest Plot for In-hospital Mortality**

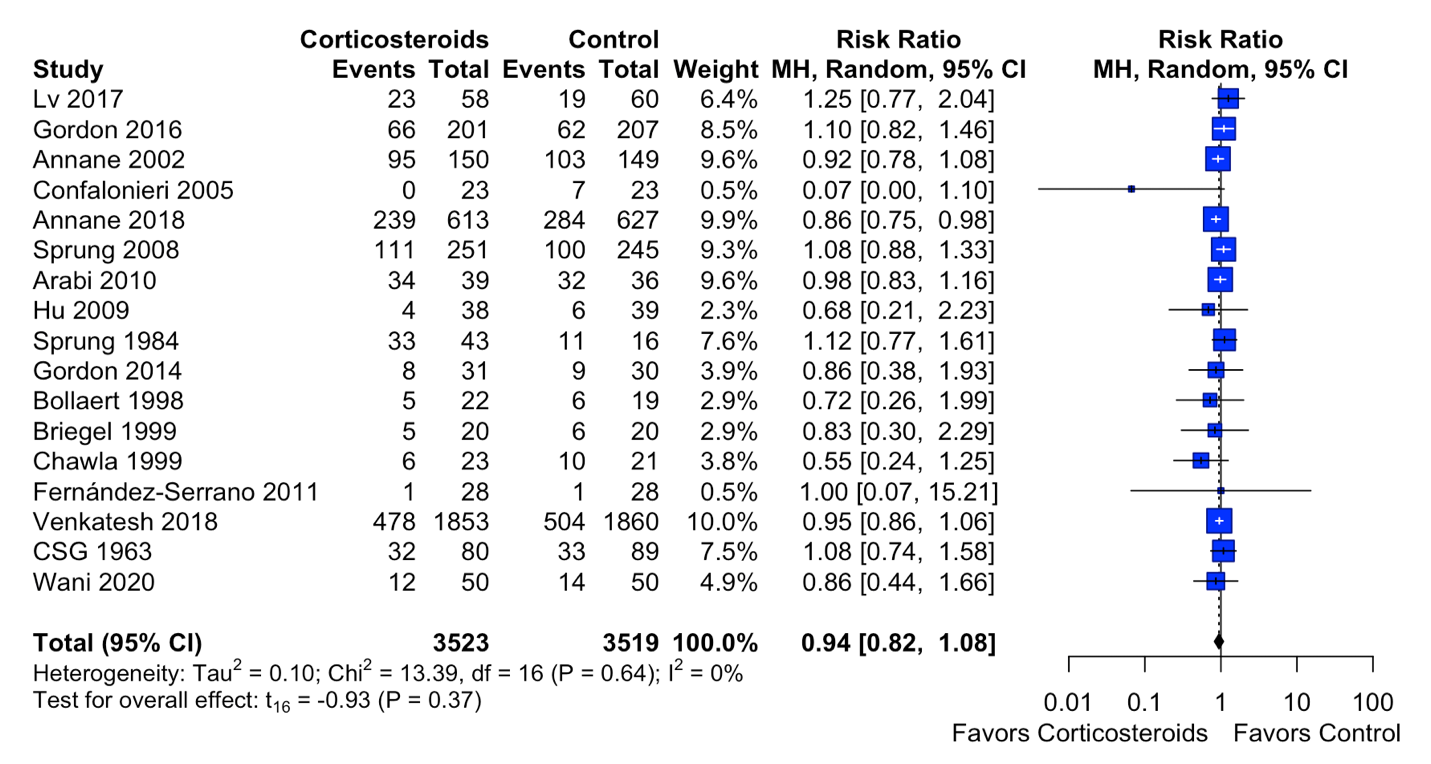

df, degrees of freedom; MH, Mantel-Haenszel method

**eFigure 16. Forest Plot for ICU Mortality**

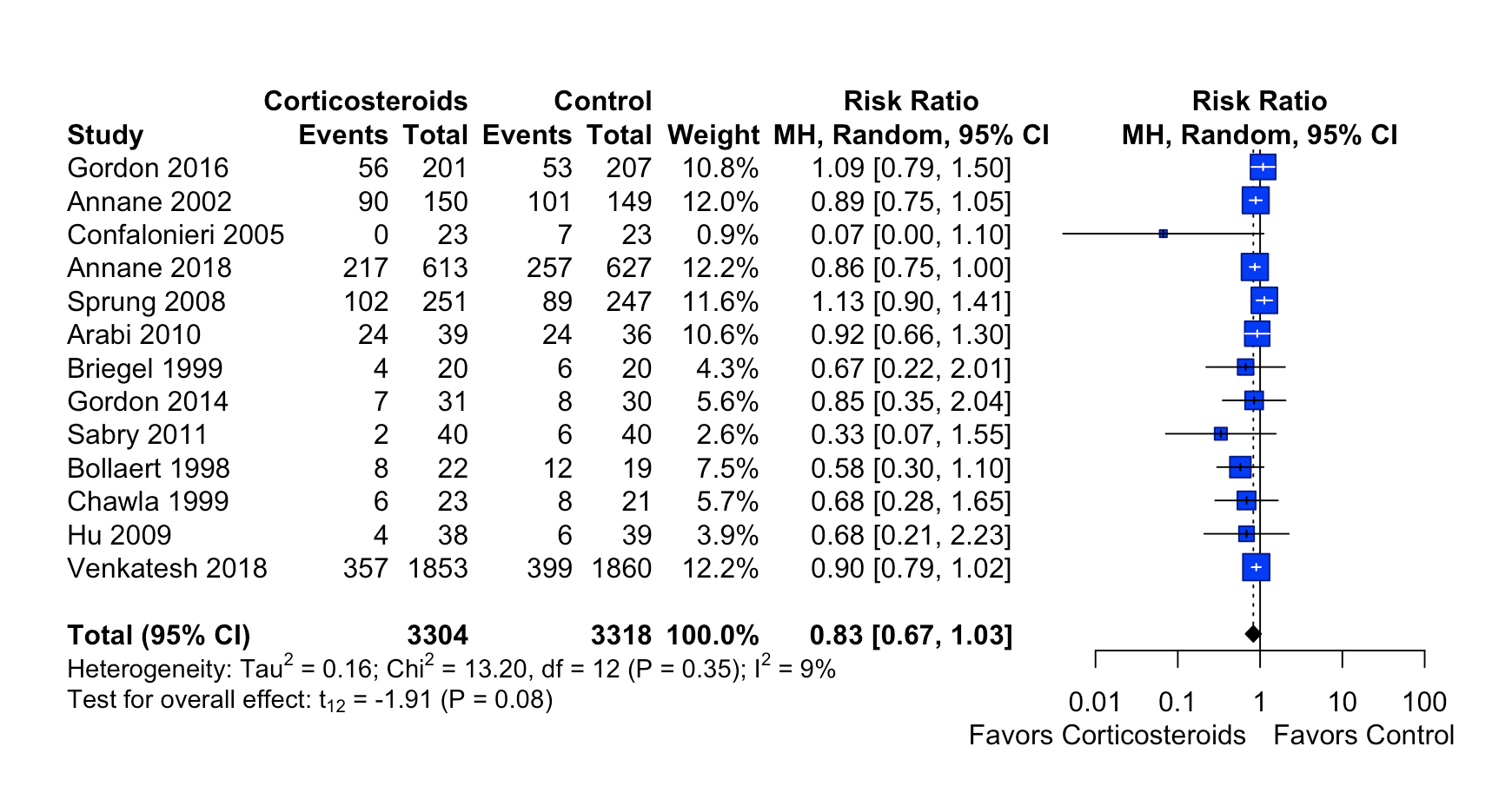

df, degrees of freedom; MH, Mantel-Haenszel method

**eFigure 17. Forest Plot for Hyperglycemia**

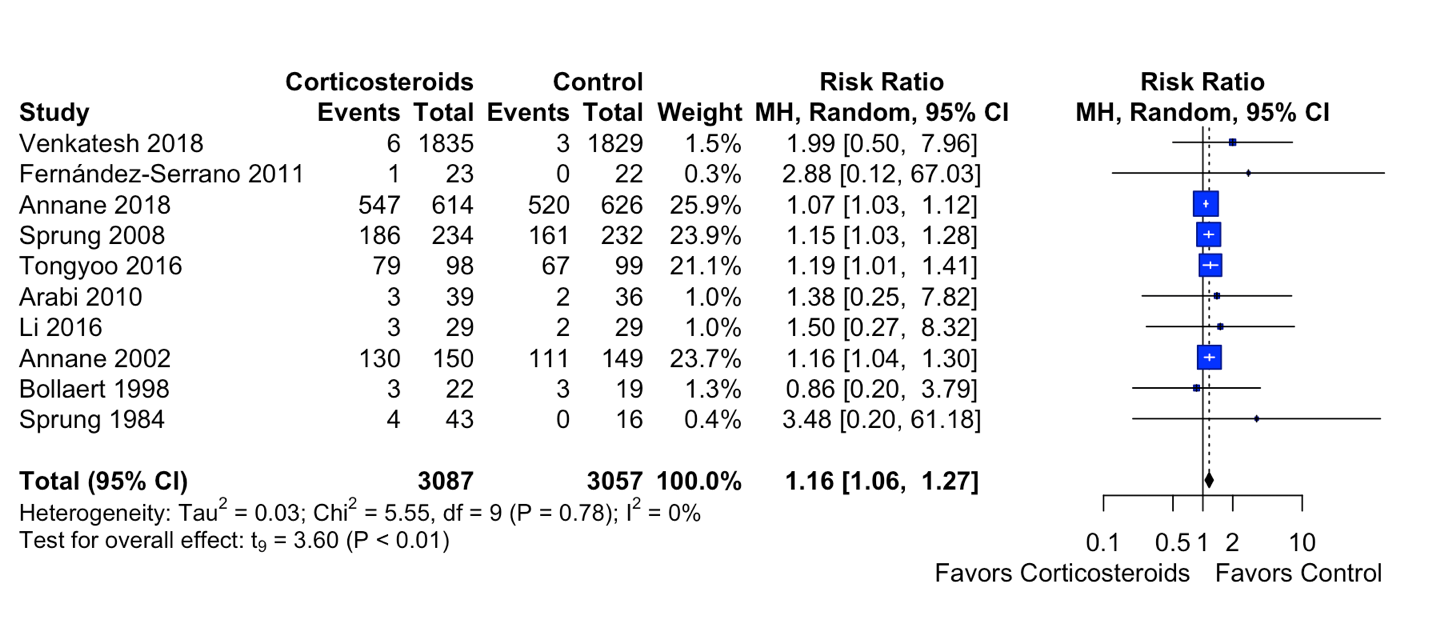

df, degrees of freedom; MH, Mantel-Haenszel method

**eFigure 18. Forest Plot for Gastrointestinal Bleeding**

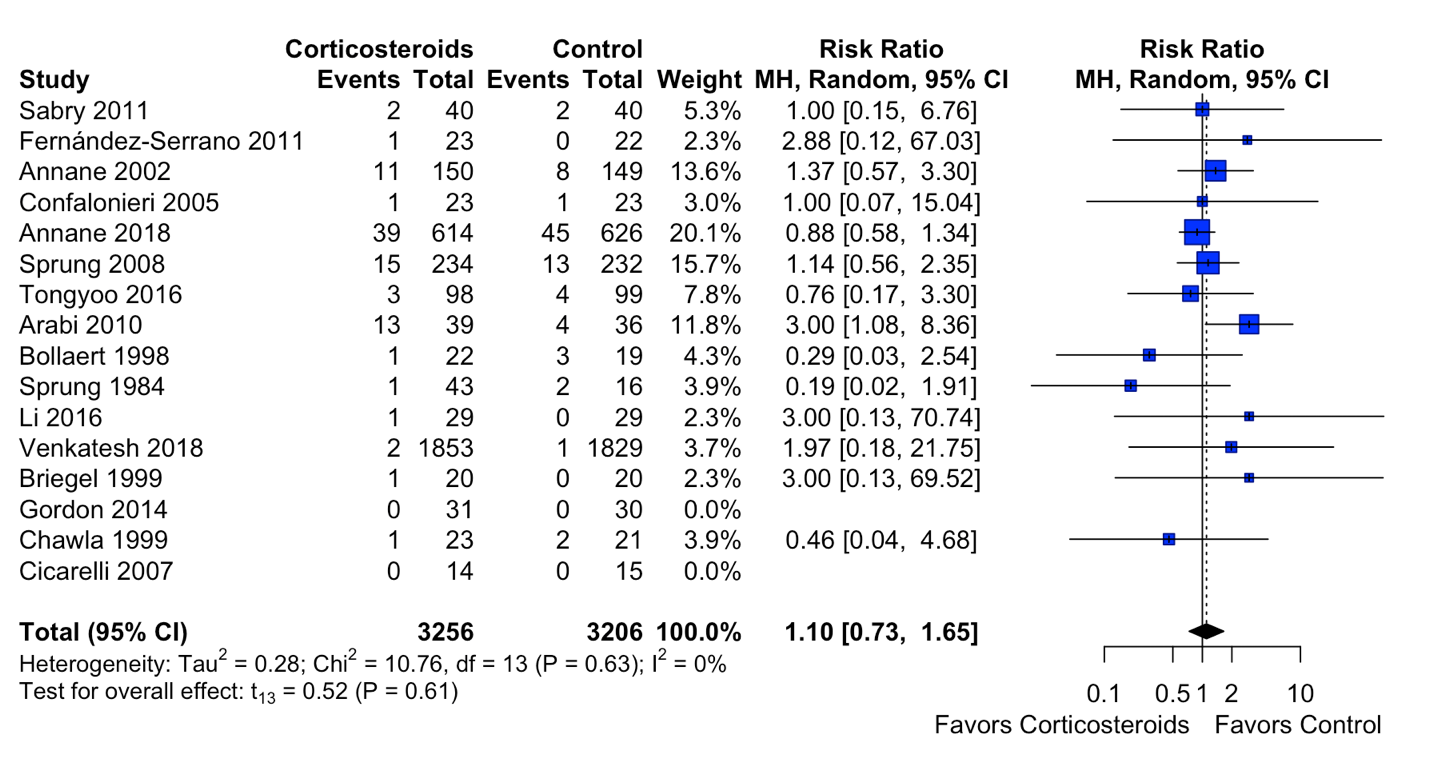

df, degrees of freedom; MH, Mantel-Haenszel method

**eFigure 19. Forest Plot for Superinfection**

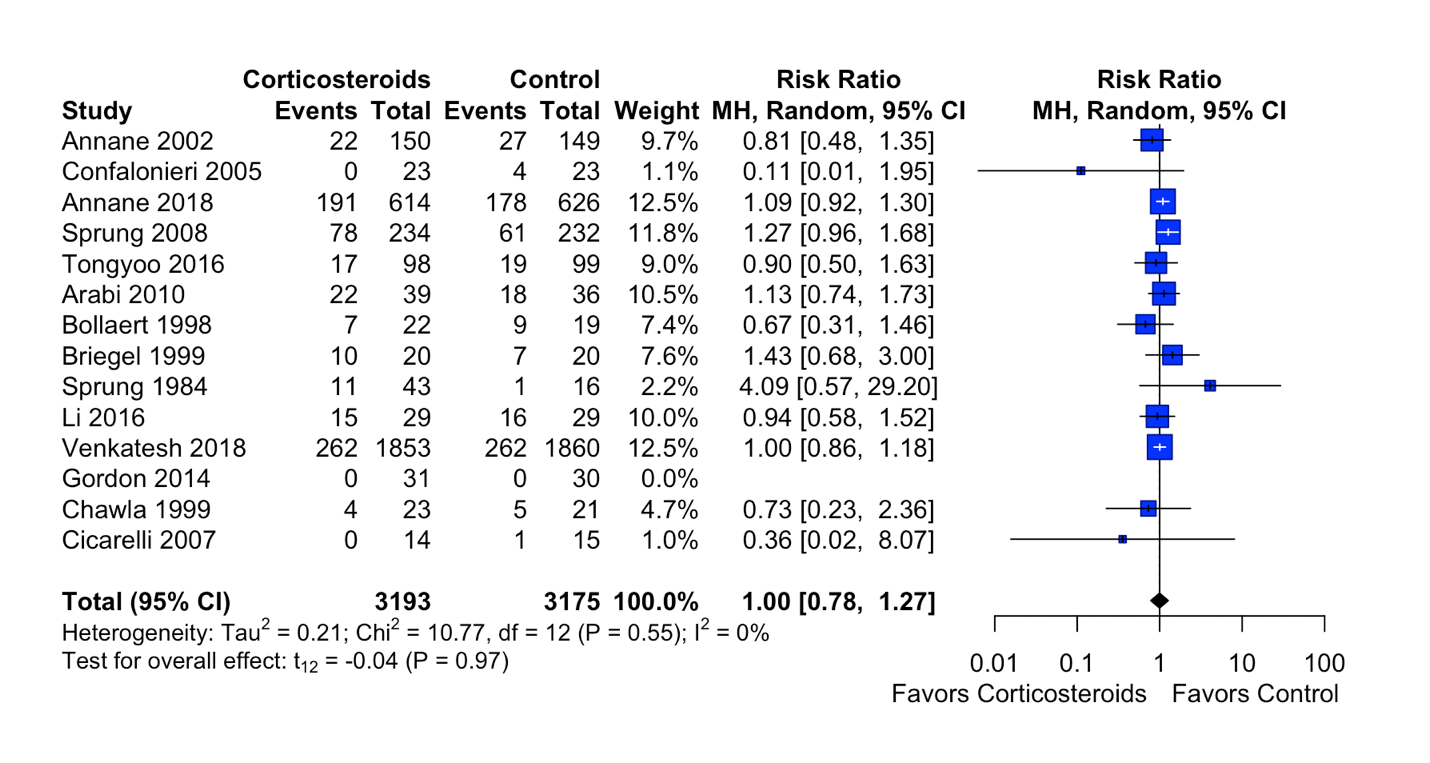

df, degrees of freedom; MH, Mantel-Haenszel method

**eFigure 20. Forest Plot for Any Adverse Event**

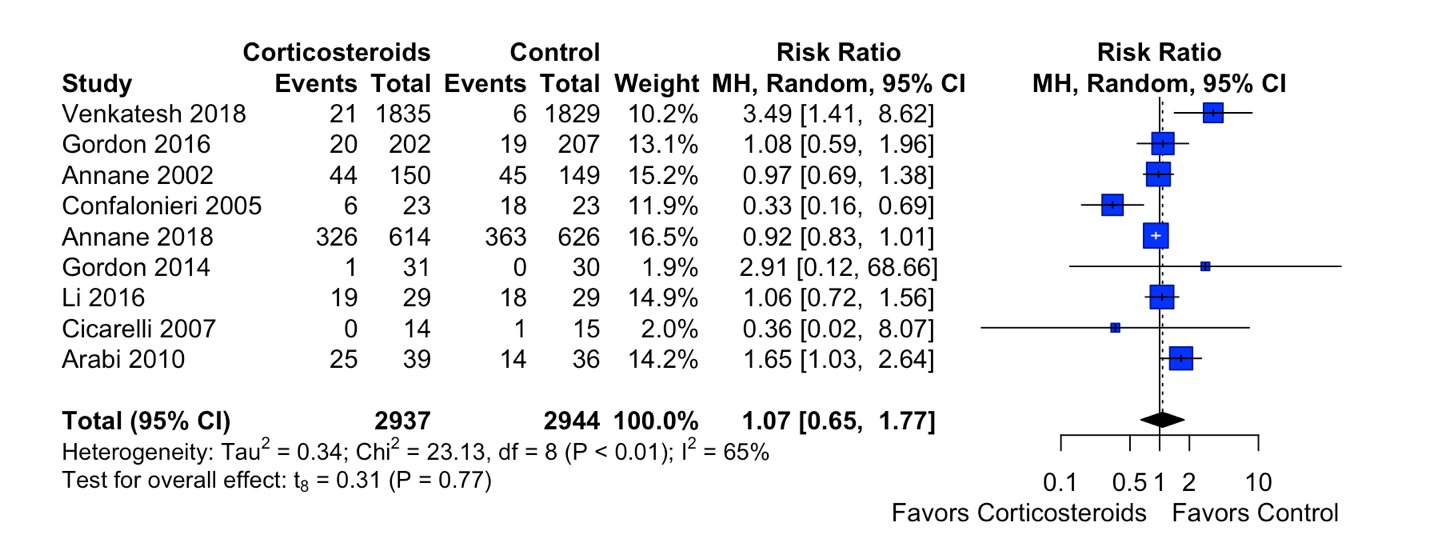

df, degrees of freedom; MH, Mantel-Haenszel method

**eFigure 21. Funnel Plot with Egger’s test for Length of ICU Stay**

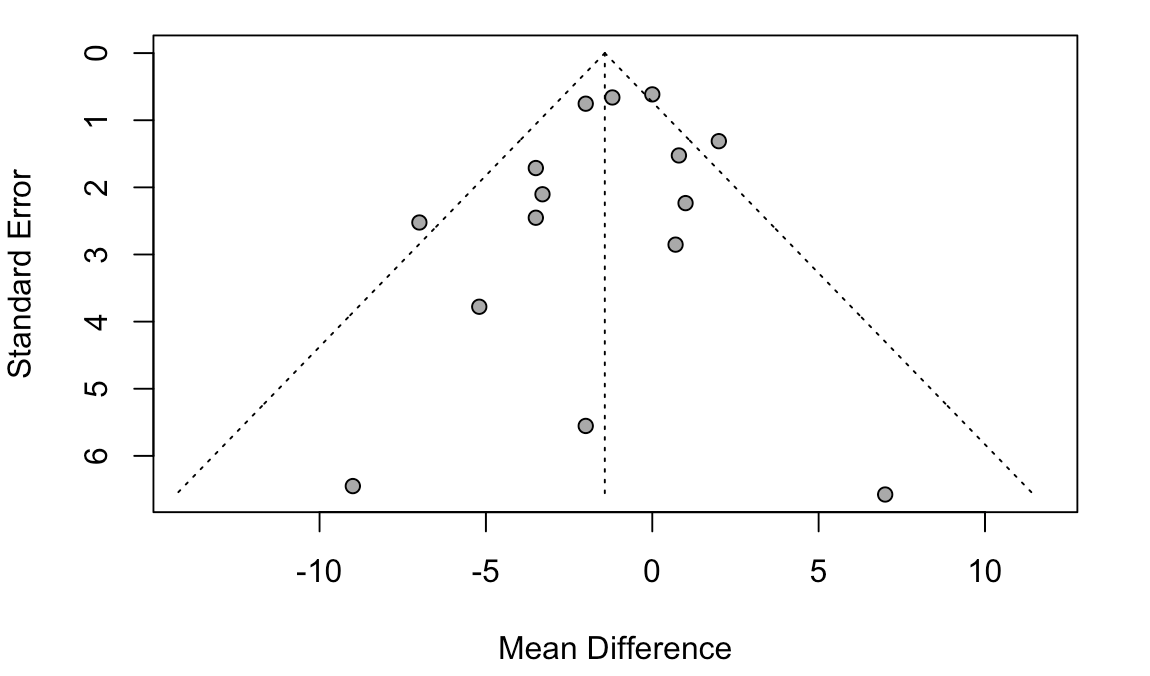

Egger's test: p=0.42

**eFigure 22. Funnel Plot with Egger’s test for Length of Hospital Stay**

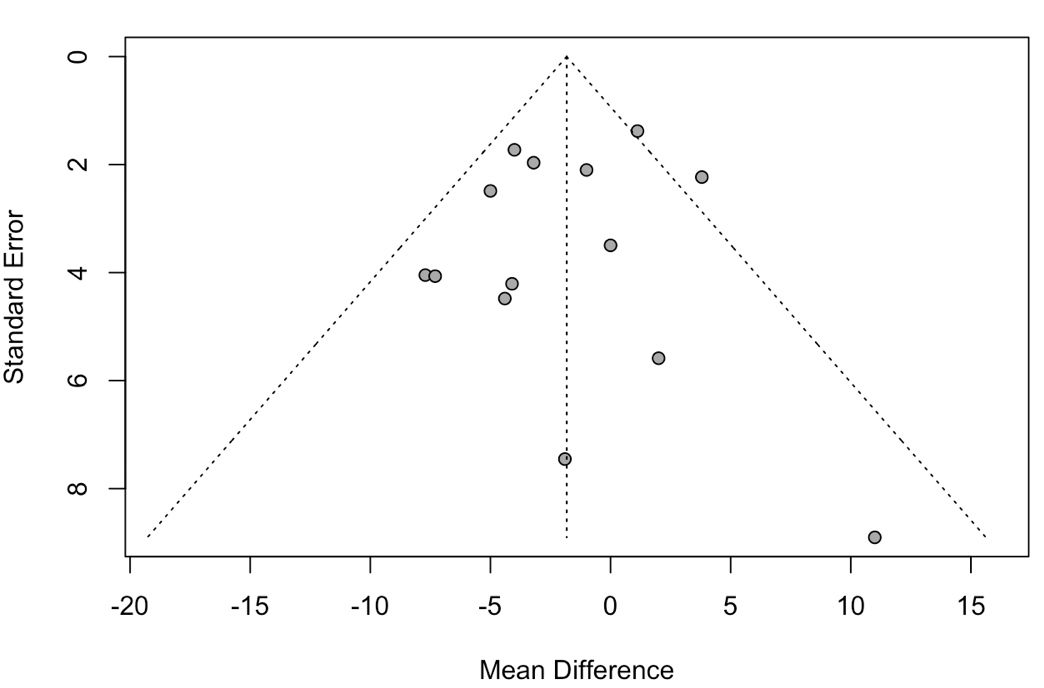

Egger’s test: 0.61

**eFigure 23. Funnel Plot for SOFA Score at Day 7**

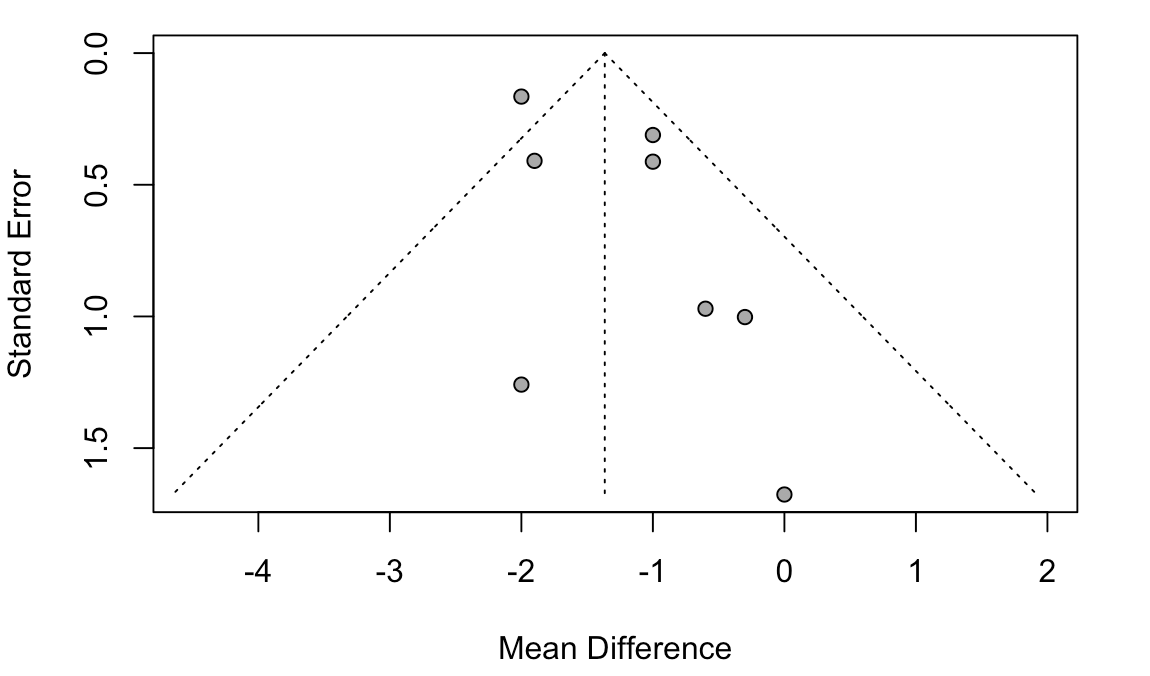

**eFigure 24. Funnel Plot for 90-day Mortality**

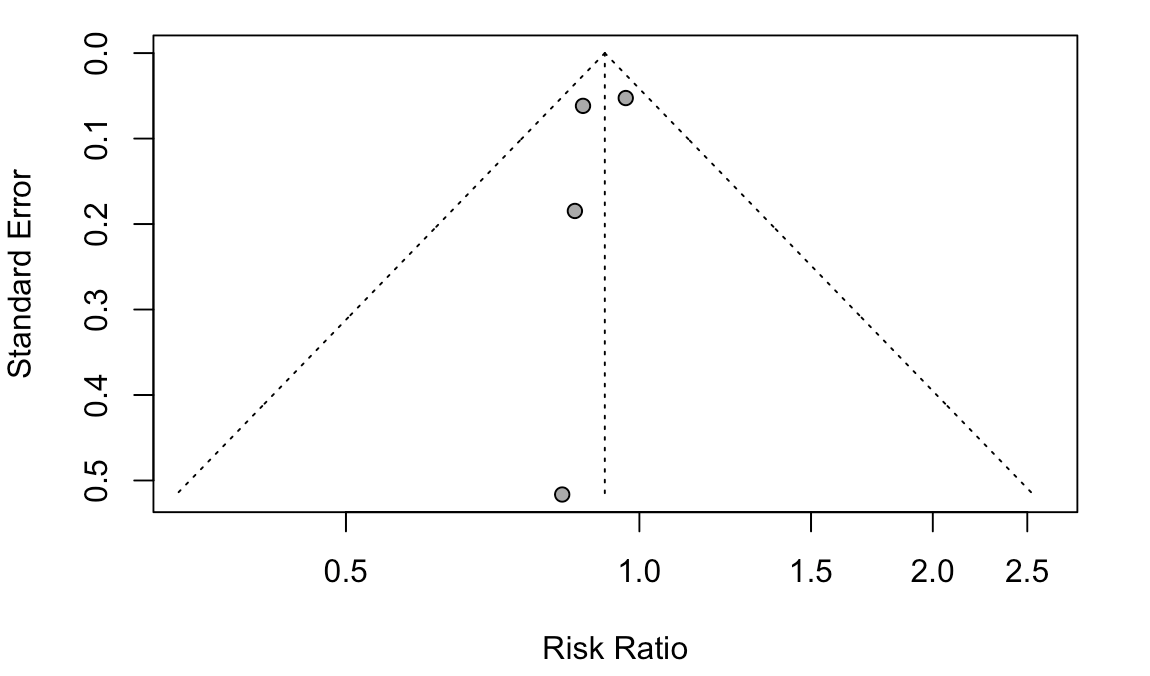

**eFigure 25. Funnel Plot with Harbord’s Test for In-hospital Mortality**

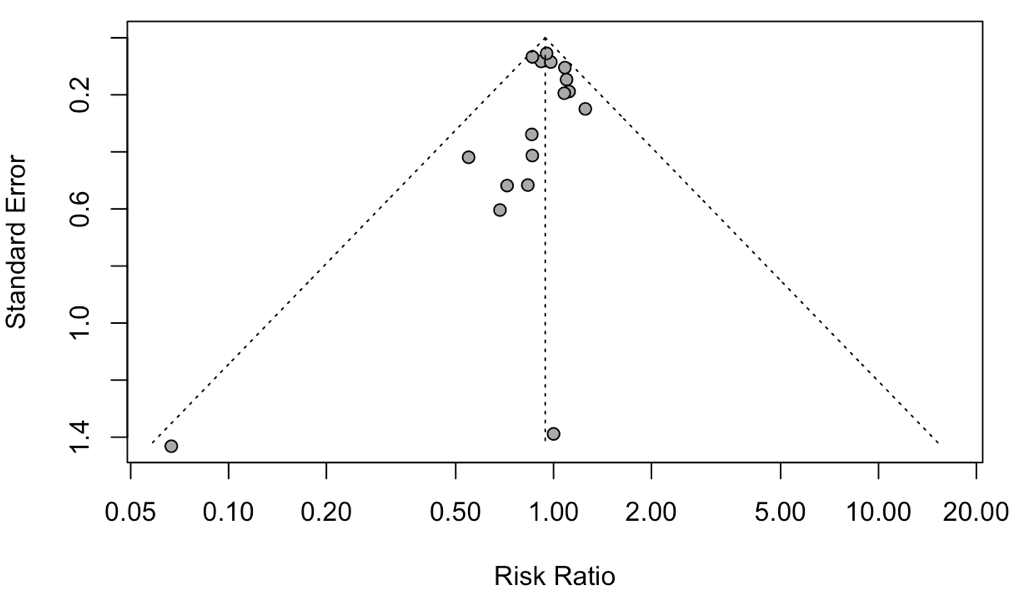

Harbord’s test: p=0.39

**eFigure 26. Funnel Plot with Harbord’s Test for ICU Mortality**

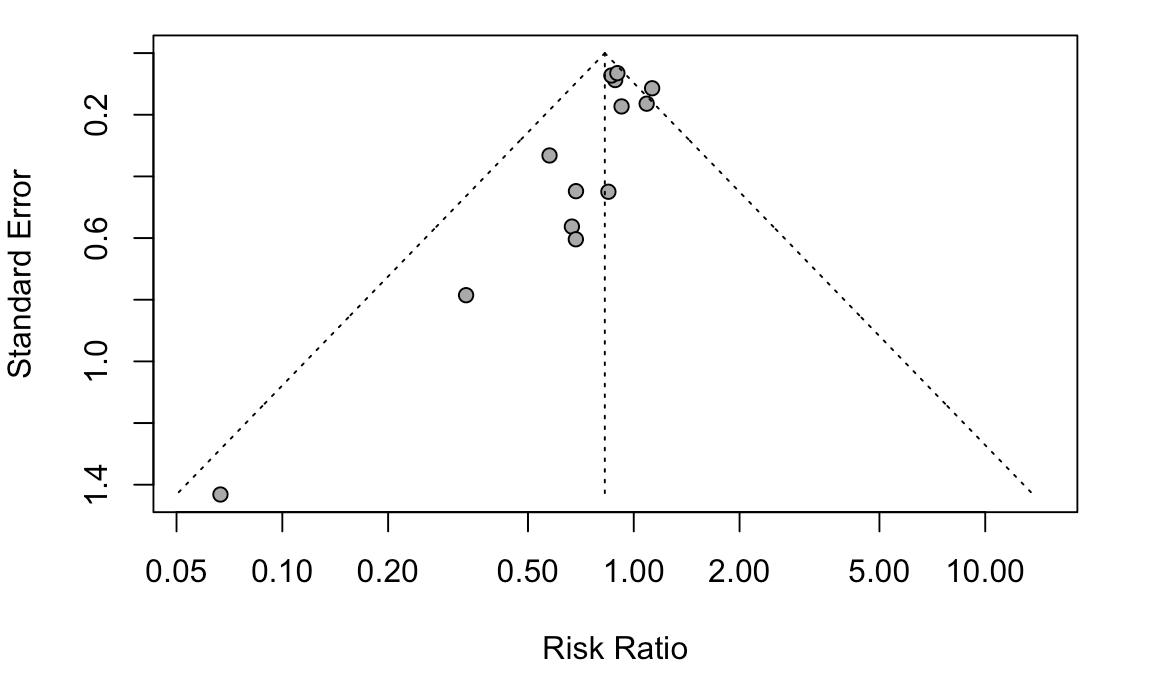

Harbord’s test: 0.08

**eFigure 27. Funnel Plot with Harbord’s Test for Hyperglycemia**

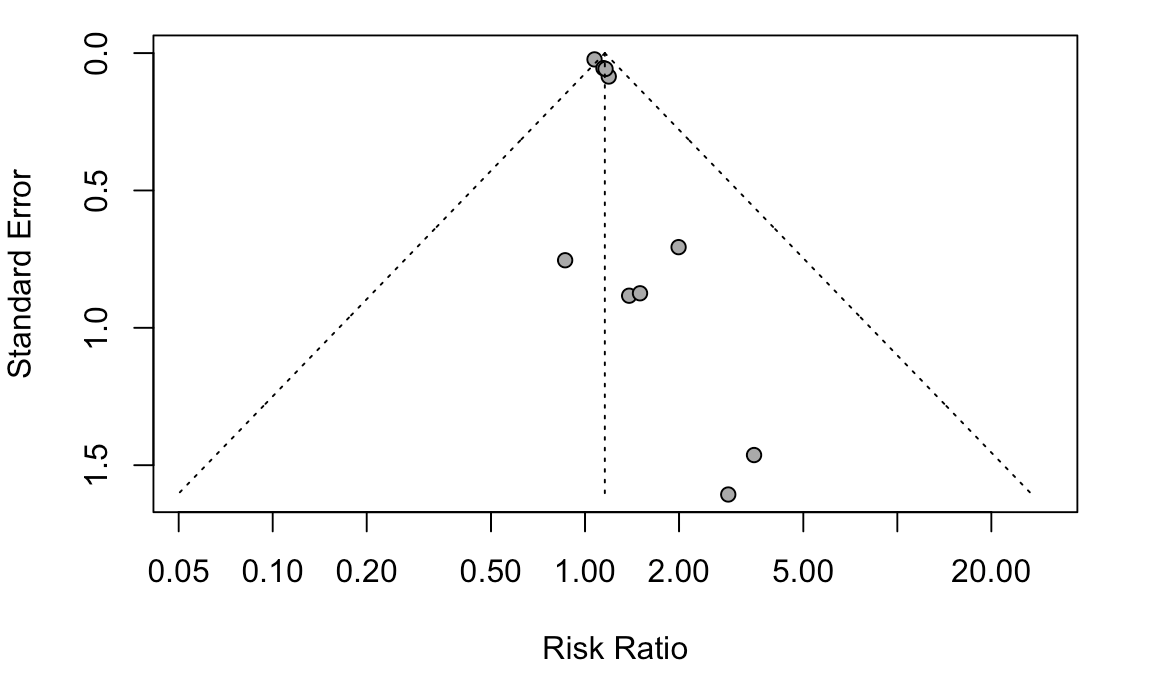

Harbord’s test: 0.02

**eFigure 28. Funnel Plot with Harbord’s Test for Gastrointestinal Bleeding**

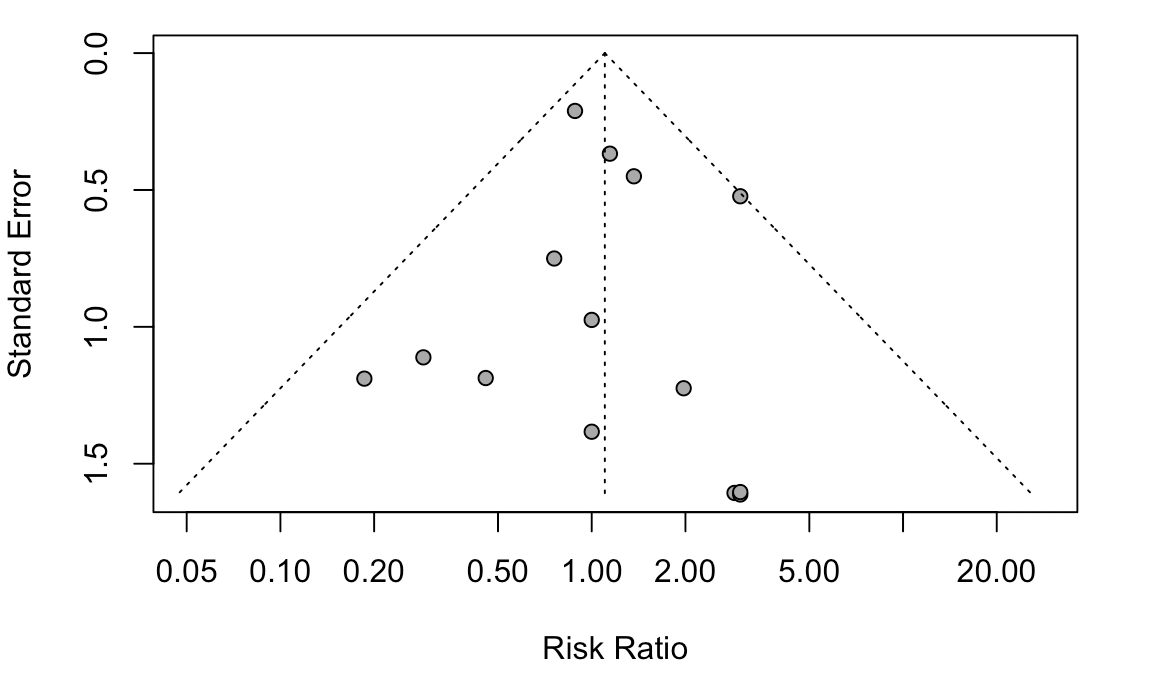

Harbord’s test: 0.63

**eFigure 29. Funnel Plot with Harbord’s Test for Superinfection**

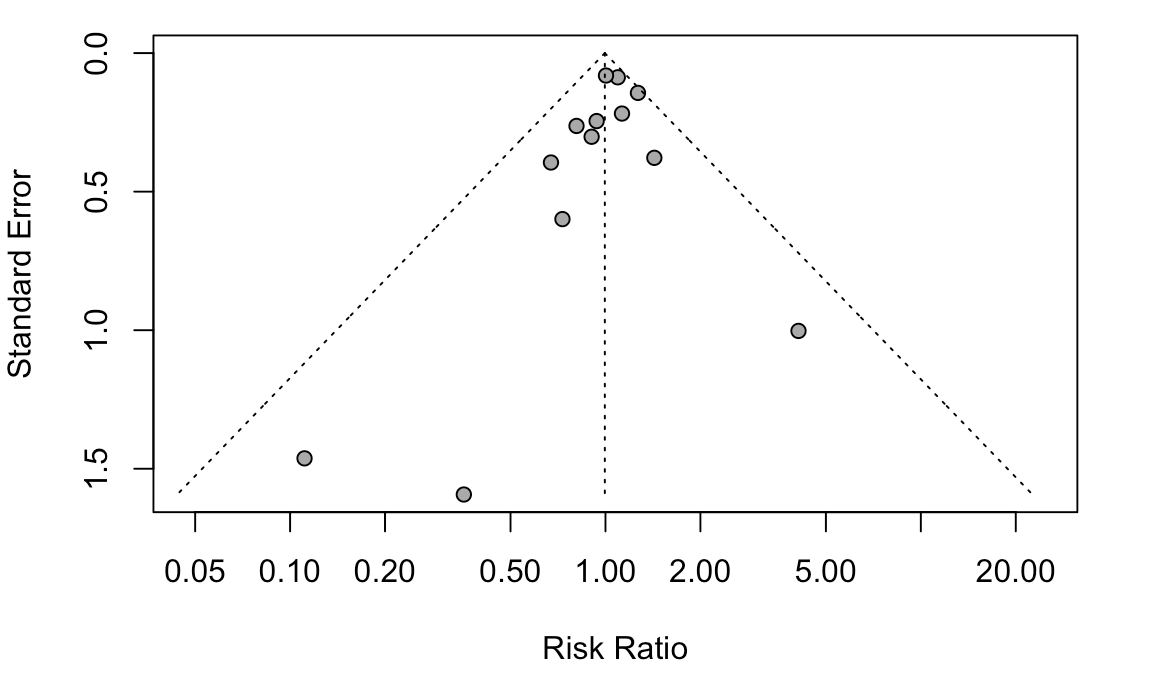

Harbord’s test: 0.21

**eFigure 30. Funnel Plot for Any Adverse Events**

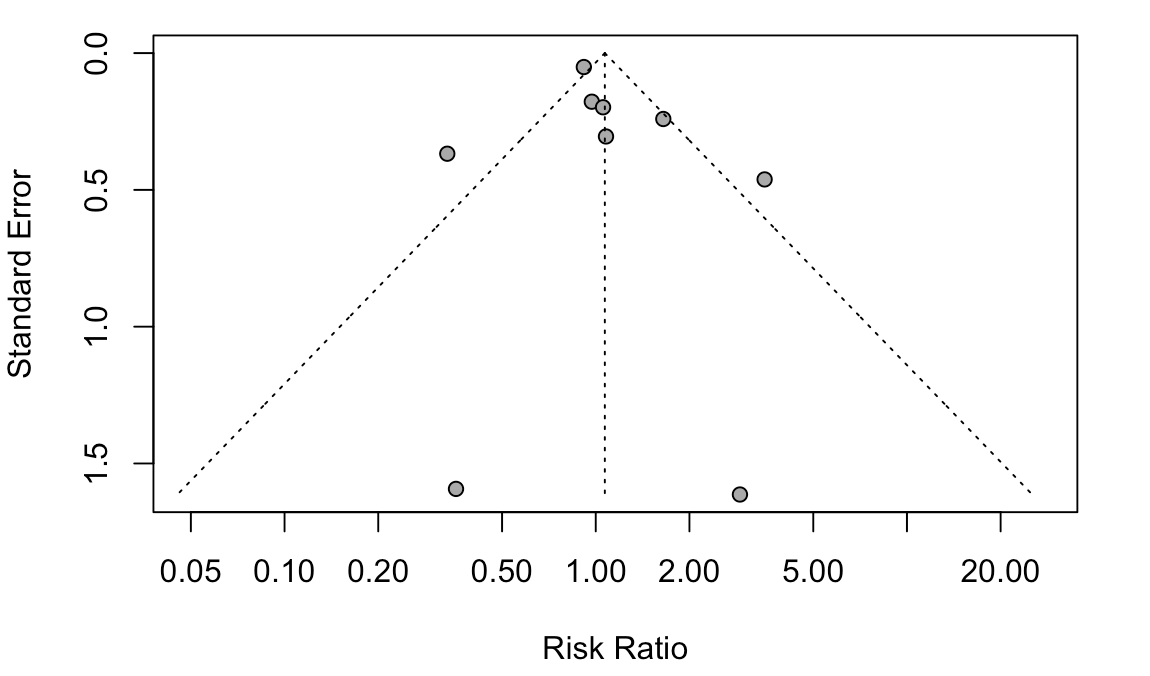

**eFigure 31. Subgroup Analysis: Stratified by Treatment Duration (**$\boldsymbol{\leq}$**7 days)**

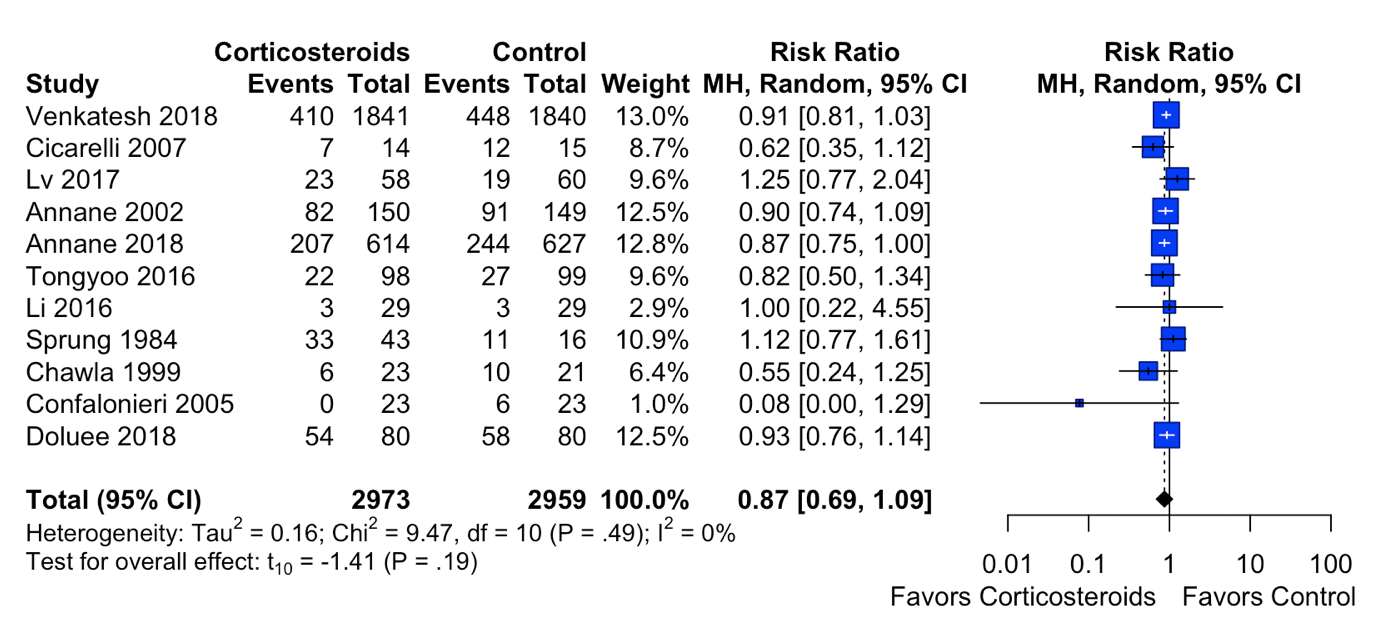

df, degrees of freedom; MH, Mantel-Haenszel method

**eFigure 32. Subgroup Analysis: Stratified by Treatment Duration (**$\mathbf{>}$**7 days)**

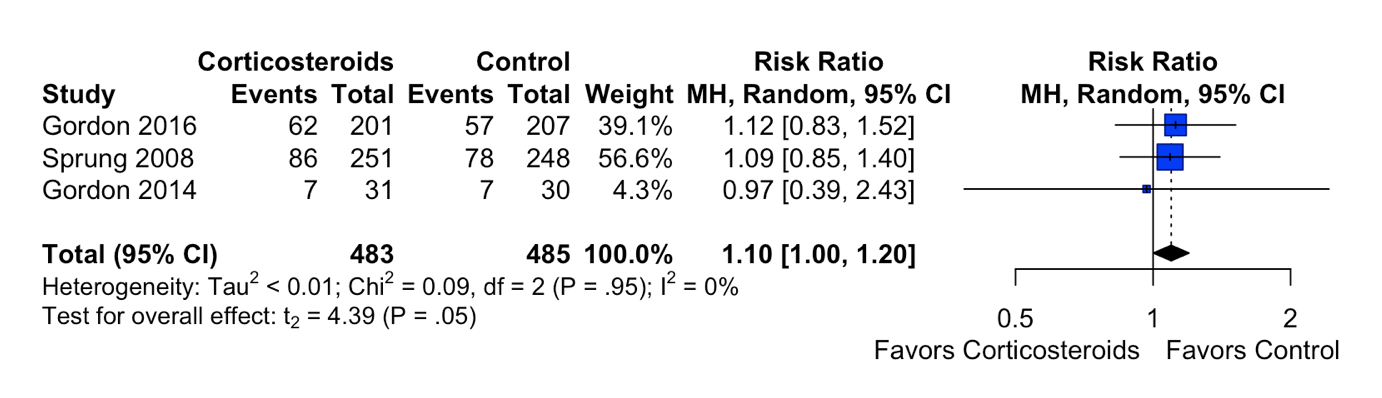

df, degrees of freedom; MH, Mantel-Haenszel method

**eFigure 33. Subgroup Analysis: Stratified by Treatment Duration (**$\boldsymbol{\leq}$**7 days vs** $\mathbf{>}$**7 days)**

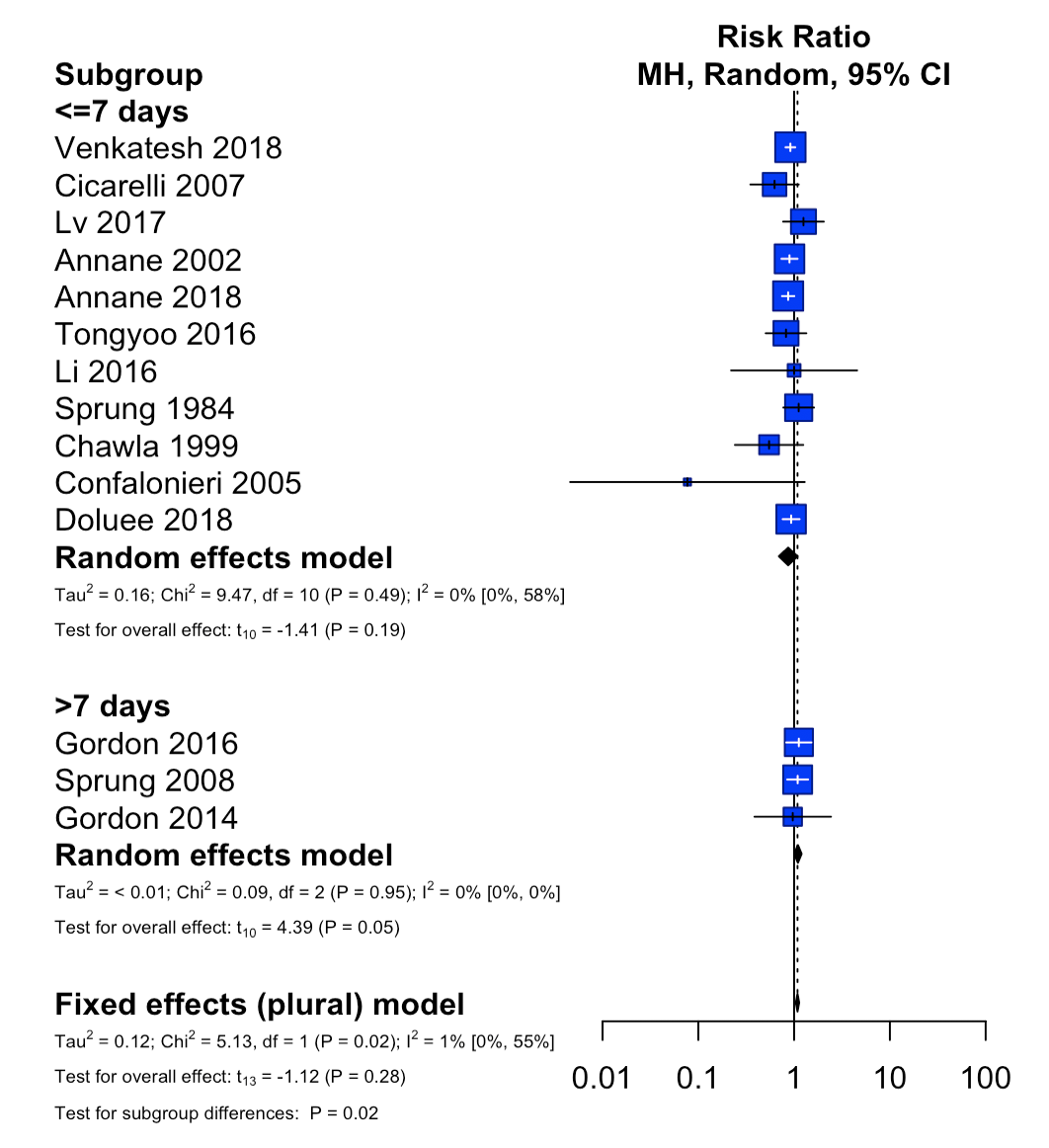

df, degrees of freedom; MH, Mantel-Haenszel method

**eFigure 34. Subgroup Analysis: Stratified by the Hydrocortisone Equivalent Dose (**$\boldsymbol{\leq}$**200mg per day)**

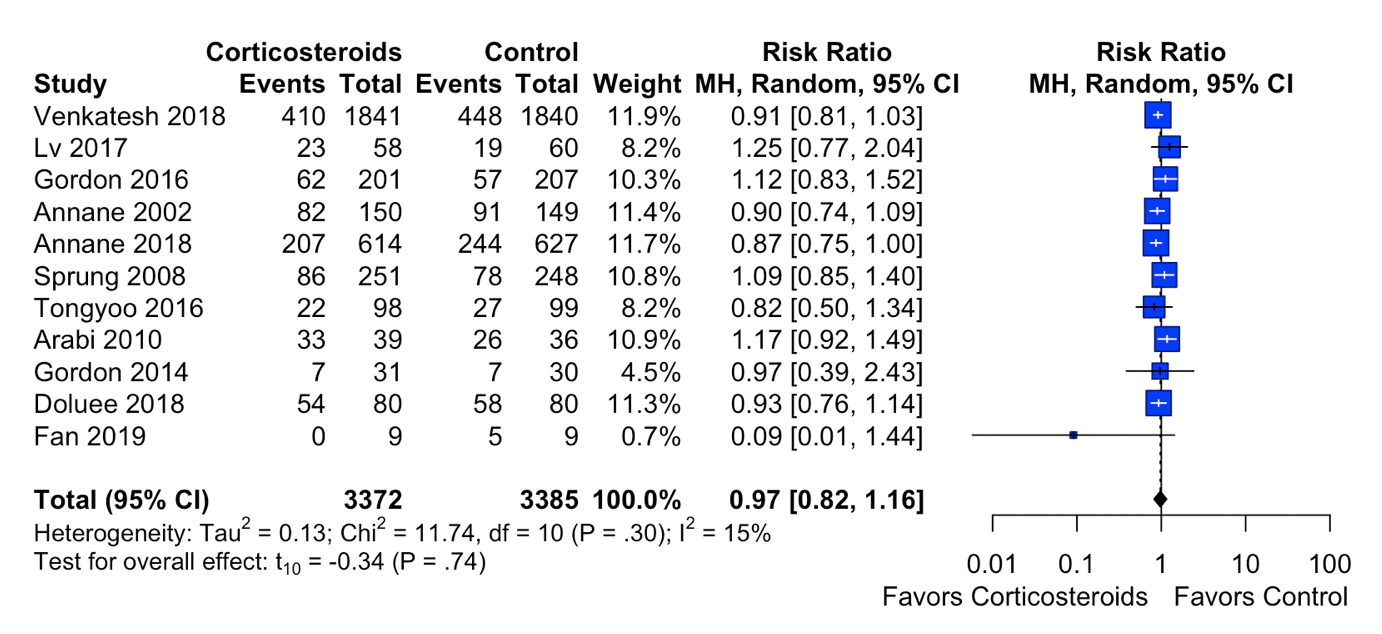

df, degrees of freedom; MH, Mantel-Haenszel method

**eFigure 35. Subgroup Analysis: Stratified by the Hydrocortisone Equivalent Dose (**$\mathbf{>}$**200mg per day)**

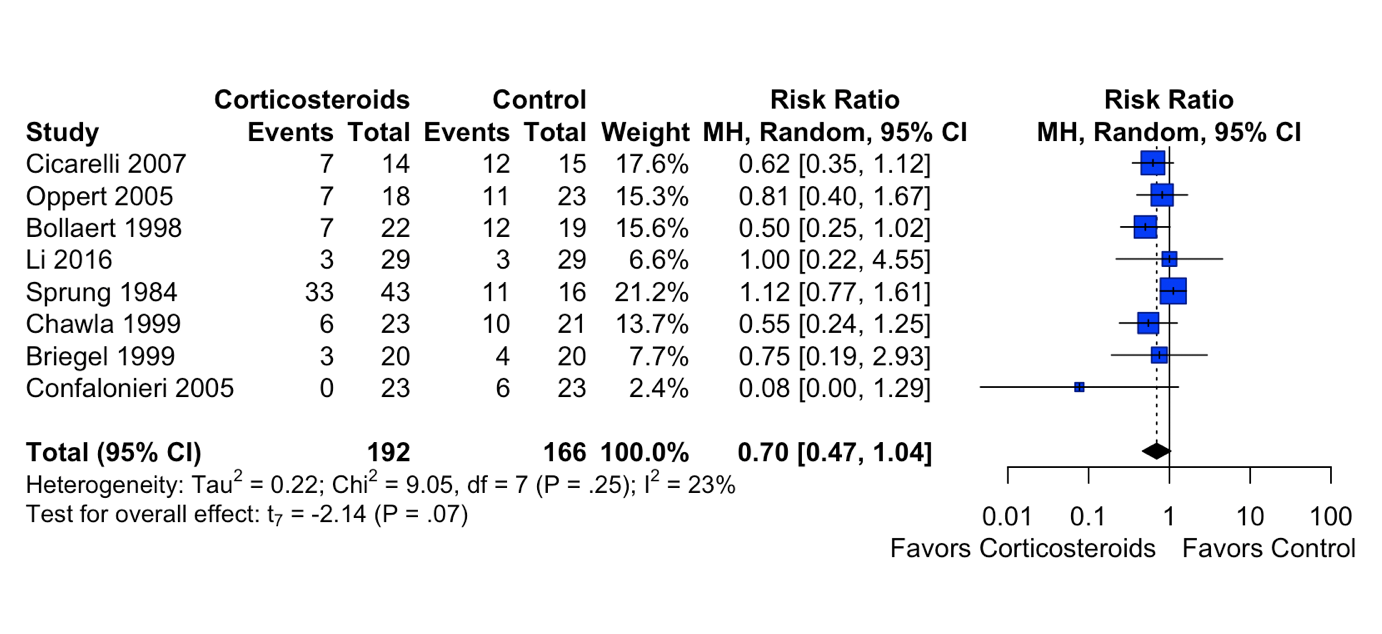

df, degrees of freedom; MH, Mantel-Haenszel method

**eFigure 36. Subgroup Analysis: Stratified by the Hydrocortisone Equivalent Dose (**$\boldsymbol{\leq}$**200mg vs >200mg per day)**

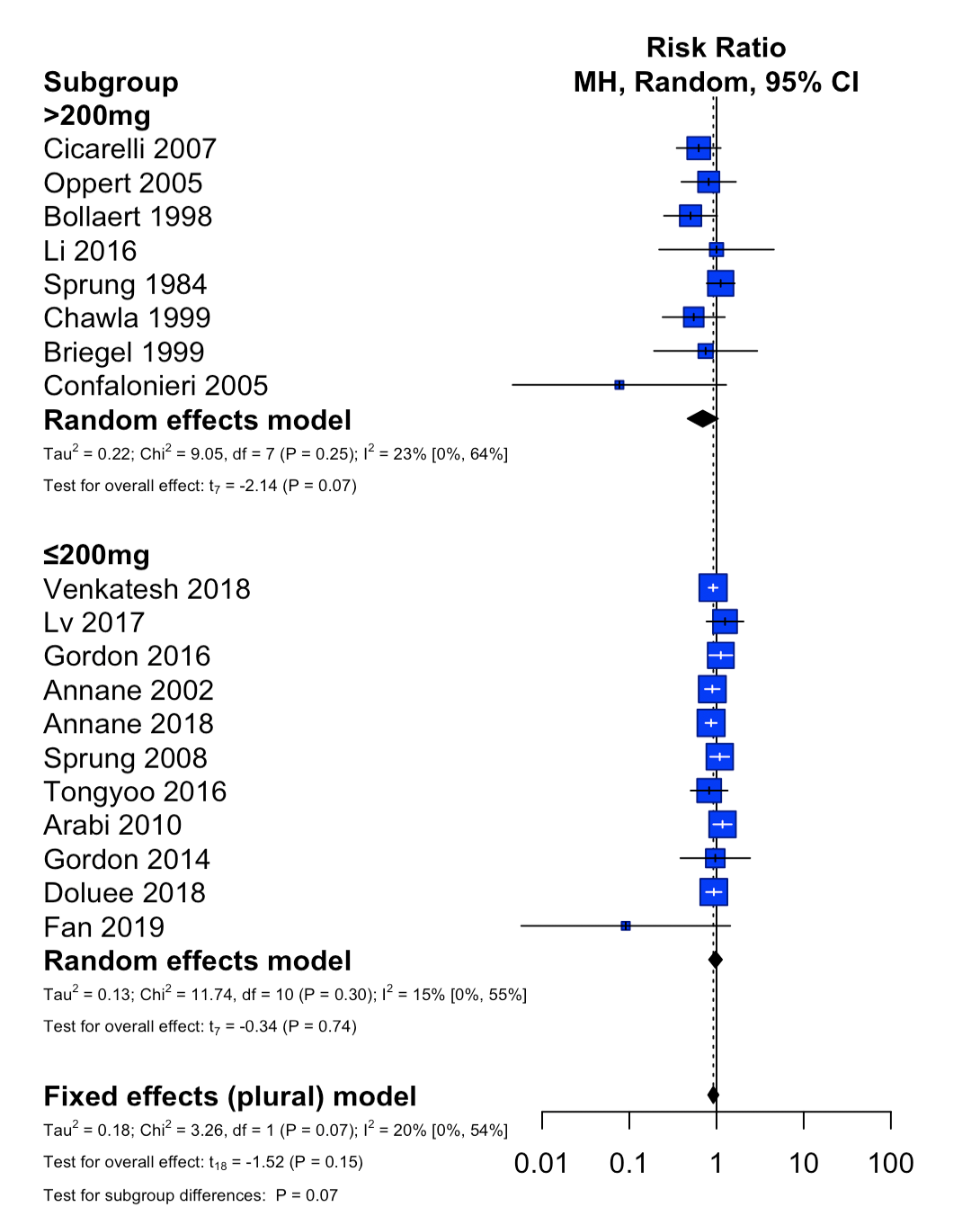

df, degrees of freedom; MH, Mantel-Haenszel method

**eFigure 37. Subgroup Analysis: Stratified by the Type of Corticosteroid (hydrocortisone)**

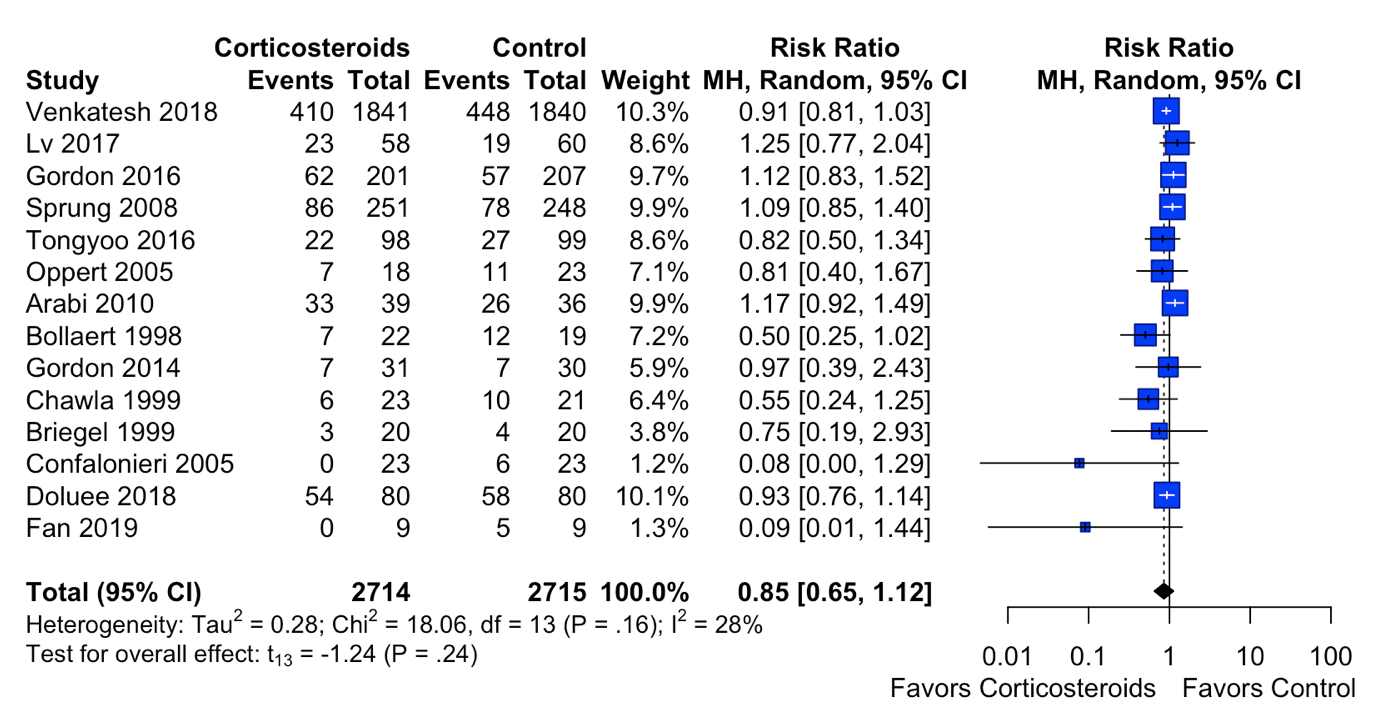

df, degrees of freedom; MH, Mantel-Haenszel method

**eFigure 38. Subgroup Analysis: Stratified by the Type of Corticosteroid (methylprednisolone)**

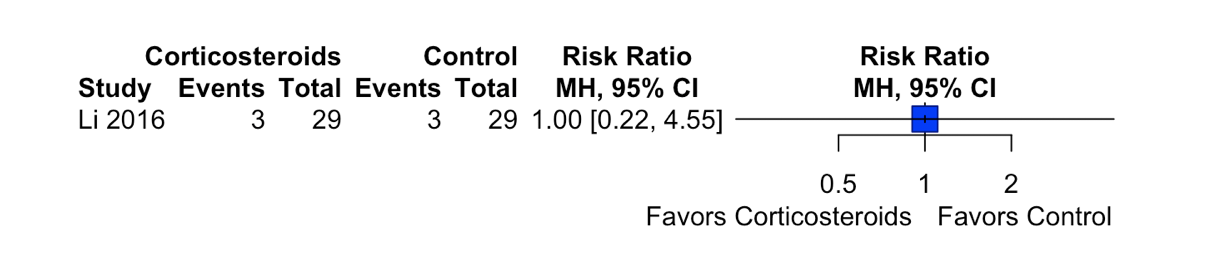
MH, Mantel-Haenszel method

**eFigure 39. Subgroup Analysis: Stratified by the Type of Corticosteroid (hydrocortisone plus fludrocortisone)**

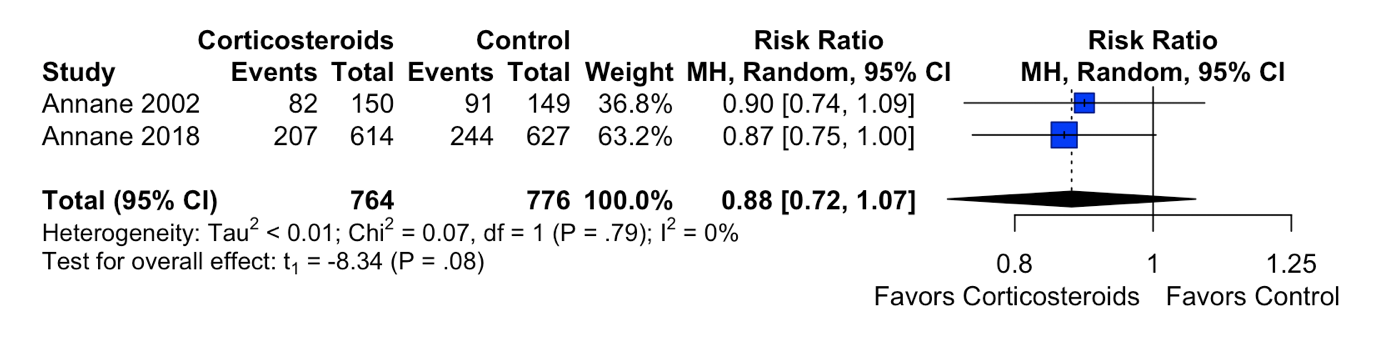

df, degrees of freedom; MH, Mantel-Haenszel method

**eFigure 40. Subgroup Analysis: Stratified by the Type of Corticosteroid (dexamethasone)**

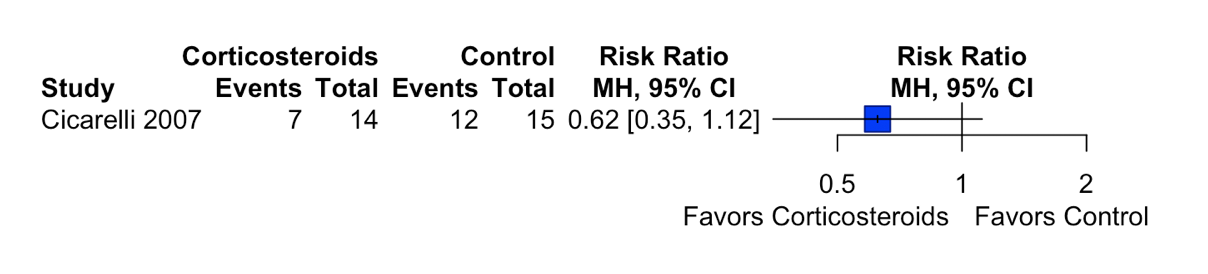

MH, Mantel-Haenszel method

**eFigure 41. Subgroup Analysis: Stratified by the Type of Corticosteroid (hydrocortisone vs methylprednisolone vs hydrocortisone plus fludrocortisone vs dexamethasone)**

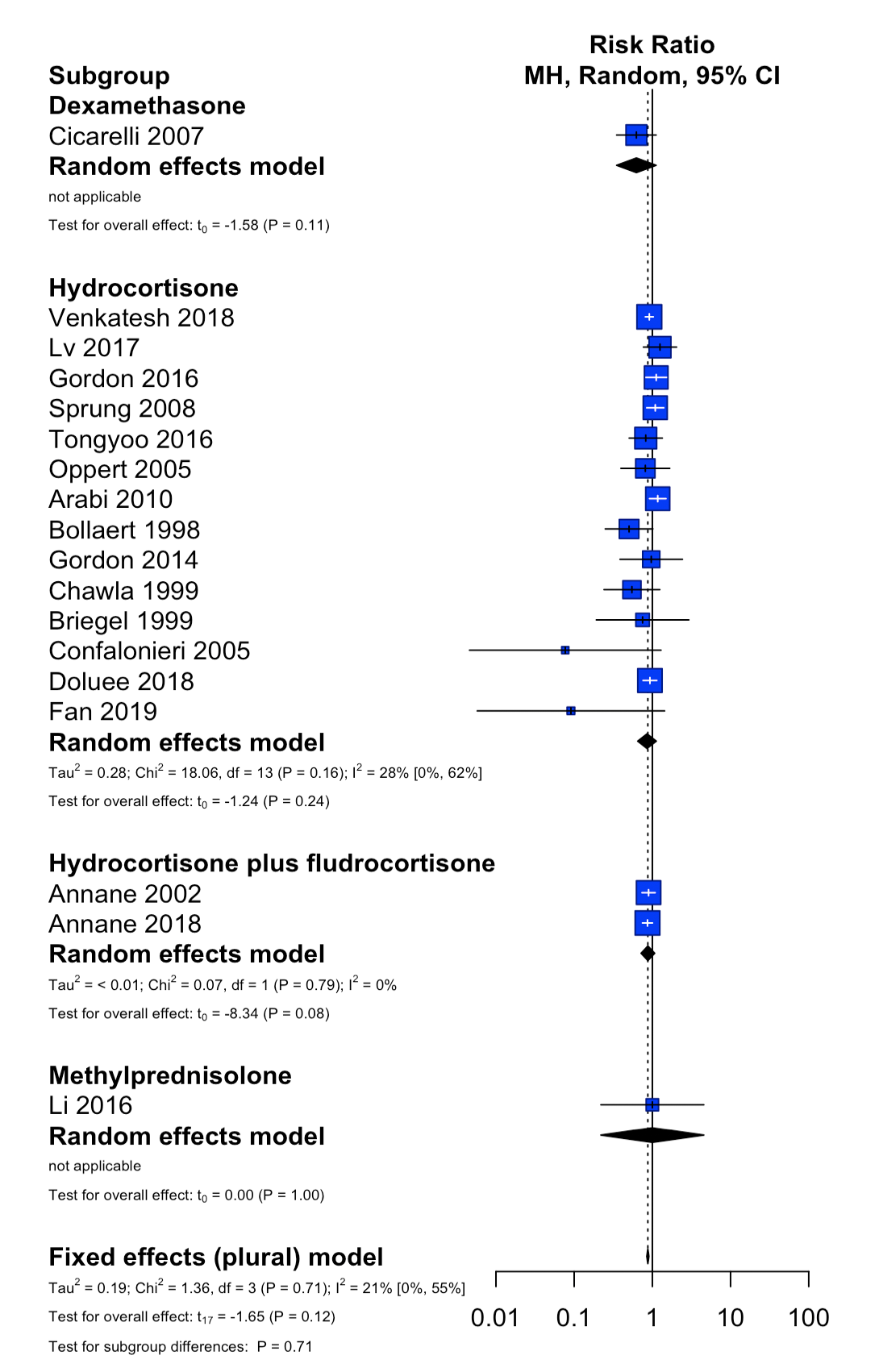

df, degrees of freedom; MH, Mantel-Haenszel method
